# Trance practice and well-being measures: the case of Auto-Induced Cognitive Trance

**DOI:** 10.64898/2026.07.15.26358125

**Authors:** Amandine Fernandez, Alexandre Foncelle, Hélène Meunier, Flavien Revillet, Audrey Breton

## Abstract

**Introduction:** Auto-Induced Cognitive Trance (AICT) is a non-ordinary state of consciousness (NOSC) that can be accessed by will alone once a standardised self-induction procedure has been learnt. The first research publication on AICT dates back only ten years, meaning that research on this phenomenon is still in its infancy. Previous reports concerning the phenomenology and neurophysiology of AICT revealed similarities with more extensively described NOSCs, as well as unusual features, raising questions about the potential benefits of AICT practice for well-being.

**Objective:** This study aimed to gather quantitative descriptive data on features associated with well-being in a large comparative sample of AICT practitioners and non-practitioners.

**Method:** This research followed a web-based survey study design which enquired AICT-trained and yet-to-be trained participants to self-report through validated standardised questionnaires on vitality, self-esteem, mental well-being, trait anxiety, life satisfaction, happiness, positive and negative affect, nature-relatedness and connectedness. Data on NOSCs practices, life history events that could have led to spontaneous NOSCs, and demographic data were collected for further inclusion as control variables in statistical models.

**Results:** The online questionnaire yielded 607 valid responses, (171 yet-to-be trained participants and 436 AICT-trained participants). AICT practice was found to be associated with increased self-esteem (RSE), overall connectedness (WCS) as well as all subdimensions of connectedness (WCS Self, WCS Others, WCS World). AICT practice Duration exhibited significant effects on global connectedness and all subdimensions of connectedness, self-esteem, trait anxiety (STAIT-5), and positive affect (PANAS+).

**Conclusions:** AICT seems to benefit to practitioners’ well-being shortly after training through increases in self-esteem and in the sense of connectedness. Prolonged AICT practice is associated with added decreased trait anxiety and increased positive affect. Further research is needed to confirm these findings with a sample including AICT-uninterested participants, and to clarify the underlying mechanisms of AICT.

## Introduction

In the last decades, non-ordinary states of consciousness (NOSCs) attained through mind-body practices, such as meditation, hypnosis, or mind-body practices (e.g. yoga, tai chi, qigong), have attracted growing scientific interest not only for what they reveal about the processes underlying human consciousness but also for their potential benefits to mental health [1–3]. Research indicates that cultivating such NOSCs through regular training can reduce anxiety and depression and improve emotional regulation[2,4,5]. NOSCs are also associated with greater self-awareness and with changes in how individuals relate to their beliefs and emotions[6,7]. These effects suggest that NOSCs may participate to enhance well-being and have likely contributed to their popularisation in contemporary societies[8] as illustrated by the rapid expansion of mindfulness-based interventions, meditation apps, and yoga programmes in contemporary Western societies.

However, while mind–body practices such as meditation and hypnosis often cultivate focused or controlled attention, or a stance of non-reactivity to spontaneously emerging thoughts, emotions and perceptions, other NOSCs arise through an opposite process of active engagement with sensations emerging from the exposure to sensory input[9,10]. The state of trance belongs to this category: it is typically induced through rhythmic auditory stimulation, repetitive movement, or other immersive sensory cues. Anthropological descriptions report a variety of induction techniques, such as drumming, chanting, clapping, or dancing, which provide an external cue that attracts focal attention on sensory perceptions and therefore facilitate entry into trance states[11–13]. Trance thus provides an opportunity to investigate an alternative, more dynamic form of NOSC and to assess how its practice may contribute to well-being.

Anthropological work has shown that in many traditional societies, trance plays a key role in healing rituals, initiation ceremonies, and mediation with spiritual entities. These functions can take two distinct but complementary forms. On one hand, “trance states” attributed to shamans, seers, channelers and traditional healers, have been traditionally associated with a state of widened/heightened perception capacity allowing the access to information otherwise imperceptible in the ordinary state of consciousness. For instance, trance experiences reported by shamans involve visions, sounds and bodily sensations perceived as otherworldly[14,15]. The acquired knowledge provides the healer/shaman with problem-solving abilities and a cosmological vision that transcends the materialistic and anthropocentric vision of life[10,16]. On the other hand, trance rituals accompanied by music or dance can be accessible to broader community members as ways to deal with emotional pain, internal struggle or re-establish personal or social harmony, as seen in Brazilian Candomblé, West African and Uyghur Sufi rituals [17–19]. Altogether, the practice of trance thus seems to be associated with changes in mindset and self-perception that facilitate problem-solving and self-regulation abilities. Such multi-cultural observations suggest that trance, while deeply rooted in ritual traditions, has universal potential to enhance well-being.

It is important to note that practices aimed at cultivating these [NOSC] states often find their origins in ancient cultural and spiritual traditions (e.g. Buddhist meditation, Hindu yogic disciplines, Taoist energy practices), which highlights their longstanding role in healing and collective support. However, the integration of NOSCs-related practices into modern Western societies often dissociates them from their spiritual anchors and redefines them instead as tools for individual well-being or even entertainment. While this disconnection can be viewed as an impoverishment, it also offers certain benefits from the perspective of experimental sciences, as it allows researchers to study specific psychological and neurophysiological effects in a controlled laboratory environment. For example, by conducting longitudinal studies on meditation training programmes, or by comparing expert and novice practitioners, studies have been able to isolate the effect of meditation on cognitive processes[20–22], quality of life[23,24], physiological response, i.e. autonomic nervous system[25,26] or cortical activity[27–29]. Despite the ubiquity of trance states across cultures, both cultural and conceptual blur may have contributed to the impeding belief that trance states are only accessible to a small number of individuals endowed with special capabilities[30], thus hindering the delineation of experimental inclusion criteria and the recruitment of participants for experimental research. As of today, scientific investigation of trance states through experimental methods is still in its infancy.

Auto-Induced Cognitive Trance (AICT) is a trance state characterised by a standardised self-induction procedure developed to make the trance state accessible to a broad population. It is designed to support scientific investigation by offering a controlled and reproducible framework. The AICT learning method relies on progressive educational milestones starting from the exposure to external inductors (sound loops) to a final stage of autonomous induction through recalling sensations associated with the trance state (voluntary cognitive engagement focused on sensations and memories). Furthermore, the AICT practice method implies building the autonomous ability to freely interrupt or sustain the trance state at will. The AICT state shares features with other more exhaustively described NOSCs such as changes in perception of time and space, in bodily sensations and sense of self, as well as emotional and mental activity. Additionally, AICT-trance is often accompanied by more specific features such as a rich mental imagery, vocal/glossolalia-type manifestations and unvoluntary movements that can be controlled with a proper training [31–33]. Contrary to the state of peaceful detachment or “non-reactivity” attained through Vipassana/mindfulness meditation[6,34], reports from AICT practitioners mention a state of intense involvement into the emerging sensorial, emotional and cognitive material, often accompanied by atypical meaning-making of the unfolding experience[32,35]. Furthermore, the experience of AICT appears both as highly subjective and variable, in that elicited perceptions are not comparable between individuals, both in their nature (auditive, visual, interoceptive, extra sensorial) and content (nature elements, objects, symbols, places, entities, memories)[33]. A recent mixed-methods study has uncovered the physiological relationship between subjective reports of AICT-trance and the adaptive ability of the parasympathetic system[36]. Taken together, this evidence suggests that AICT constitutes a dynamic process of immersion in perceptions influenced by the individual and their state, which may indicate a state of heightened self-awareness.

At another level, data on AICT phenomenology reveals experiences relating to the individual’s relationship with their environment. These experiences show similarities to advanced meditative states and psychedelic-induced experiences. For instance, AICT has been associated with experiences of self-dissolution and loss of bodily boundaries[37–39] as observed in meditative practices[40] and under the influence of DMT or Psilocybin psychedelics[41–43]. As with classic psychedelic-induced trances, relationship with the environment during AICT can manifest as dialogue or non-verbal interaction with symbolic or natural presences, or transformations into natural features or non-human living beings[33,44]. Interestingly, both an increased awareness of self-related processes and a sense of environmental connectedness have been identified as key mechanisms involved into mental-health improvements produced by psychedelic-assisted therapy[43]. The existing parallels between AICT phenomenology and NOSCs associated with mental-health benefits strongly support the investigation of AICT’s effects on well-being. Three mechanisms highlighted by AICT research offer insight into how this NOSC may influence physiological and psychological dynamics, either transiently or lastingly. Firstly, changes in ANS activity observed during the AICT state suggest modulation of the nervous system towards decreased parasympathetic activity. This has been interpreted by the authors as engagement of a vagal self-regulation mechanism[36,45]. Secondly, retrospective data on AICT experiences have shown an association between a transient state of selfless psychological functioning and increased happiness during the AICT state[39]. Thirdly, decreased EEG synergistic activities in delta, alpha and theta bands have been observed during the AICT state, which authors interpreted as reflecting a reduced external awareness and modified sense of self observed during the AICT state[29]. AICT may therefore act as a powerful catalyst for transforming self-experience and environmental relationship.

The enhancement of mental, emotional and physical flexibility in order to achieve a state of inner peace, clarity of mind, emotional balance and harmony with oneself and the external world lies at the core of the principles of yoga and meditation [46]. The hope conveyed by these traditional approaches to well-being and their associated meditative practices has since been supported by research into traditional and secular adaptations of these practices. A meta-analysis documented the positive effects of mindfulness meditation-based practices on reducing emotional exhaustion and depersonalisation and increasing personal accomplishment in the burnout-vulnerable population of healthcare professionals[47]. Other studies on meditation and mindfulness-based interventions have reported improvements in attentional control[27] and cognitive flexibility[21]. One of the core features targeted by meditative practices, i.e. mindfulness, has been identified as having a pivotal influence in favour of emotional adjustment and emotional well-being[48] and to act as an enhancer of happiness through the mediation of improved emotional stability and self-esteem[49]. Improvements in mindfulness were also observed following yoga interventions, which were further associated with benefits including interrupting negative thinking, reducing depressive symptoms, and alleviating stress and anxiety (see[50,51] for a review).

The exploration of NOSCs potential on the improvement of well-being is also being tackled through the ongoing revival of research on psychedelic substances targeting mental-health issues. Psilocybin in particular, has been associated with breakthroughs in patients concerned by major depression and treatment-resistant depression[52,53]. Qualitative analysis of patient reports undergoing psilocybin-assisted therapy highlighted the efficiency of psilocybin on favouring emotional acceptance and inducing a state of “connectedness”, perceived by patients as pivotal on their recovery [43,54]. The same report of transitioning from a state of separateness to interconnectedness while experiencing a dose of psilocybin was also observed in patients experiencing a high state of anxiety associated with a cancer diagnosis[55]. While a systematic review identified connectedness as a recovery process in mental health[56], the benefits of a sense of connectedness were also supported by research involving non-clinical populations on hair cortisol [57], psychological well-being, meaningfulness and vitality[58]. Overall, research on the science of well-being, whether seeking its underlying factors through fundamental science, or involving variously induced NOSCs, highlights influential features including - but not limited to-emotional stability, self-esteem, and a multifaceted sense of connectedness. The current study aims to evaluate whether well-being itself and/or the aforementioned factors may benefit from training in, and sustained practice of, the secular NOSC practice of AICT.

The standardised method provided by AICT practice offers a unique opportunity to investigate a homogeneous, trance-type NOSC in a population of practitioners. The descriptive data on the phenomenological and physiological changes elicited by this NOSC portrays a unique combination of features observed in behaviourally and substance-induced NOSCs, which suggests possible benefits on various dimensions of well-being. This study sought to investigate which possible dimensions of well-being may be influenced by AICT practice and prolonged practice. Because of the multilayered nature of the well-being construct, our objective was to gain insight into both subcomponents of well-being and underlying processes favouring this state through a combination of standardised scales. Our approach thus included both classic and newly designed validated psychometric tools targeting the well-being subcomponents of vitality, happiness, life satisfaction, positive and negative affect, anxiety, and mental well-being, as well as the well-being-supporting processes of connectedness, nature-relatedness and self-esteem. Because current research data lacks a comprehensive description of widespread AICT effects, we opted for an online survey design to maximise reach among AICT practitioners. We also aimed to identify which dimensions of well-being may benefit from immediate effects of AICT practice, and which may require prolonged practice to arise, with respect to influence of life events and mind-body practices susceptible to induce confounding NOSCs episodes. Our sample openly included participants with any amount of AICT practice, to enable time-dependent comparisons. Participants were enquired about situations at risk for-(i.e. coma episode), and occurrences of spontaneous trance episodes, as well as side practices of other NOSCs to control for confounding effects. Our design further includes a population composed of AICT-untrained participants whose interest for AICT was comparable.

## Methods

### Participants

The collected data were cleaned according to validity criteria prior to statistical analysis. The validity criteria used and the corresponding data output are described in **Figure 1** below. Out of 1004 initial responses to the online questionnaire, 397 were excluded, yielding 607 valid responses—171 from untrained participants and 436 from AICT-trained participants. Using the demographic data, we computed an index of individual socioeconomic level according to the method of Genoud [59]. AICT practitioners refer to participants who have received the introductory four-day AICT practice workshop delivered by the TranceLab Training Institute, which teaches them how to induce, maintain and cease the state of AICT trance at will and autonomously. These participants will be referred to interchangeably as ‘AICT practitioners’ or ‘AICT-trained’ throughout this paper. The untrained participants were people on the waiting list for a training session. All participants aged 18 years and above, formerly trained or yet to be trained to AICT, not pregnant nor concerned by a legal protection, a known neurological condition or psychiatry diagnosis were eligible to participate. Despite the five-month open enrolment period and continuous new subscribers joining the training organisation’s waiting list, the number of untrained participants could not reach that of the already trained participants (over 3000). Therefore, the amount of data collected on trained participants exceeded that collected on untrained participants.

**Figure 1.**
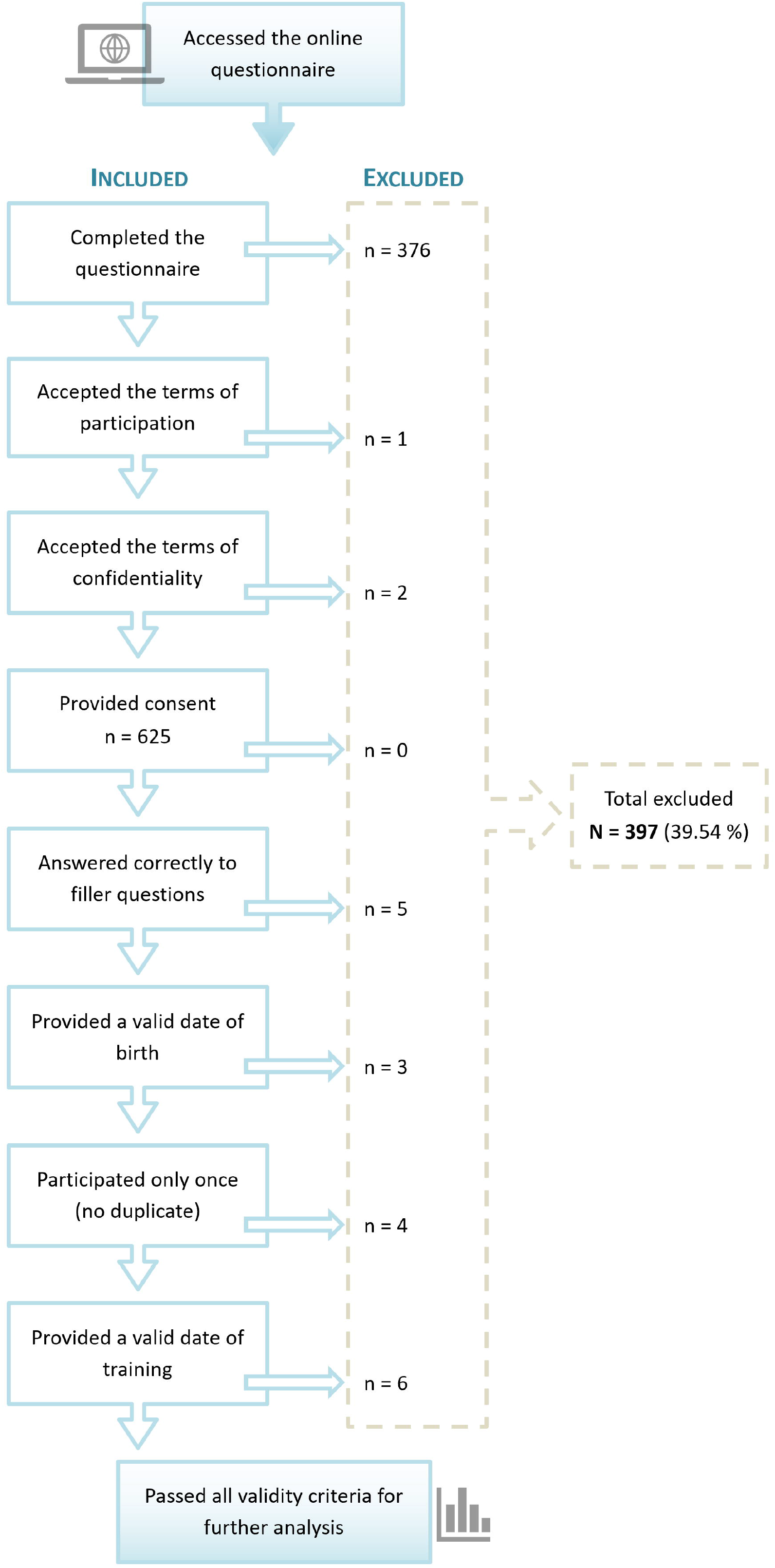
Flow chart description of validity criteria applied for data cleaning and resulting outcomes.

### Study design & procedures

The call for participants was disseminated through an email campaign forwarded by the AICT practice organism, TranceLab Training Institute (https://trancelabinstitute.com/), and displayed online on the platform “Trancers’ Lab”, a platform only accessible to AICT trainees following the completion of their training workshop. The email campaign and online display on “Trancers’ Lab” allowed for the advertisement of yet-to-be trained and formerly trained participants that would constitute untrained, and AICT-trained populations, respectively. The advertisement of both AICT-trained and untrained participants through the channels of the AICT training organism intended to control for mind openness towards trance-related subjects so as to minimise implication and self-evaluation biases. This research was carried out using an anonymous online survey elaborated through the LimeSurvey client (https://www.limesurvey.org/fr). The survey was kept online and available to participants from February 23^rd^ 2024 to June 3 2024. The announcement briefly described the purpose of the research study, altogether with the list of inclusion and exclusion criteria, the names of the investigators, and estimated duration of the survey. The interested participant was invited to click on a hyperlink to access the online survey if found eligible. The welcome page detailed the participant’s rights, research objectives, risks and benefits. Participants were informed that their enrolment was voluntary, and that answers were collected anonymously. Participants were invited to confirm that they had read and understood the eligibility criteria and to tick a box indicating whether they believed themselves to fulfil the criteria. Participants accessed successively to 9 validated standardised scales in their full or abridged form in a random order. All scales were used in their French version or in-house French-translated in the absence of an existing French version. Once completed, participants were guided through a series of questions asking about their practices of NOSCs, life history of events that could have led to spontaneous NOSCs, and demographic data. Collection of these data allowed for their further inclusion as control variables in statistical models. Two filler questions were positioned at an estimated one-third and two-thirds of participants’ progression through standardised questionnaires. These consisted in two easy multiple-choice enquiries designed to assess participants’ attentional involvement in fulfilling the questionnaire (“How much is 10 + 12?”; “Which word does not make reference to an animal?”). The questionnaire lasted approximately 20 to 35 minutes. No incentives were offered to participants of this study.

### Sample-size

In addition to the dependent variable of AICT practice, our data analysis included a multivariate analysis covering nine covariates (age, gender, socioeconomic level index, autohypnosis practice, meditation practice, relaxation practice, trauma experience, endangered life experience, spontaneous trance experience). According to the statistical recommendation for multivariate analysis of a minimal 10 to 20 entries per independent variable [60,61], a minimum cohort of 180 participants was required to achieve statistical power. To meet this requirement and given the current lack of quantitative data on the well-being of AICT practitioners, our target was set at a minimum of 500 complete survey responses.

### Questionnaires

- **STAIT-5**: this scale is a five-item short form of the Spielberger’s State-Trait Anxiety Inventory designed for the evaluation of trait components of anxiety, whose psychometric properties were shown to be comparable to that of the full-form (Cronbach’s alpha for STAIT-5 = 0.82)[62]. Respondents are invited to rate five statements related to the experience of anxiety on a four-point Likert scale (1 – Not at all; 2 – Somewhat; 3 – Moderately so; 4 – Very much so) according to their experience in everyday life. The highest score of 20 describes the highest level of anxiety. In the absence of a validated French version of the STAIT-5, we relied on the translation of the corresponding items from the French validated full-form STAI[63], by taking reference on the STAIT-5 items described in references [62,64].
- **SHS-F**: a validated French version [65] of the Subjective Happiness Scale [66] in which participants are asked to self-evaluate through four statements using a seven-point Likert scale. The overall subjective happiness index is obtained by averaging the four scores, with the global score of seven indicating the highest level of happiness. The SHS-F has been positively correlated with mindfulness and self-compassion and was shown good internal consistency (Cronbach’s alpha = 0.83)[65].
- **SVS**: the Subjective Vitality Scale was originally designed as a seven items scale allowing respondents to self-evaluate their psychological and physical vitality using a seven degrees Likert scale (from 1 – Not at all true; to 7 – Very true)[67]. Subsequent structural equation modelling by Bostic et al. [68] highlighted a better fitting model of the scale as a measure of vitality when removing item n°2 (negatively worded). In this study, we used a French translation of this six-item scale version whose internal consistency (Cronbach’s alpha = 0.83) was shown to be consistent with Bostic’s et al. [69]. The items were formulated according to the “individual difference level” version of the scale, in which participants are asked to rate how true each statement is for them in general. According to the French SVS, the overall score of each respondent was obtained by summing the six items’ scores.
- **RSE**: the Rosenberg Self-Esteem Scale is a ten-item scale related to feelings of self-esteem, self-competence, and self-acceptance recently considered as the most popular psychometric tool for the assessment of self-esteem [70]. Although a two-dimensional structure explained by the scale’s content of positively and negatively worded statements has been suggested, the one-dimensionality of the scale has been supported by the prominence of its general factor (self-esteem) in explaining the variance of the items [71]. In this study, we used a shortened eight-items version of the Canadian French validated version of the RSE. The psychometric properties of the Canadian French version of the RSE were shown to be comparable to that of the original scale (Cronbach’s alpha between 0.70 and 0.88 across four studies; test-retest correlation r = 0.84 for the Canadian French RSE) [72]. Respondents were invited to rate their agreement to the eight chosen statements using a four degrees Likert scale (1 – Strongly agree, 2 – Agree, 3 – Disagree, 4 – Strongly disagree), a higher total score being indicative of a higher self-esteem.
- **SWL**: the Satisfaction With Life Scale allows for the self-assessment of global life satisfaction through five statements with a seven-point Likert scale (1 – Strongly disagree, 2 – Disagree, 3 – Slightly disagree, 4 – neither agree nor disagree, 5 – Slightly agree, 6 – Agree, 7 – Strongly agree). The scores of the items are summed and give an overall score comprised between 5 and 35. A higher score therefore pictures a higher degree of satisfaction with life [73]. This scale was shown to be unifactorial and have high internal consistency both in its American original form and in the Canadian French (Cronbach’s alpha = 0.80) validated version used for the purpose of this study [74].
- **WEMWBS**: the Warwick Edinburgh Mental Well-Being Scale is a mental health related scale consisting of 14 positively worded items covering hedonic and eudaimonic aspects of mental health [75]. Respondents are requested to indicate the frequency of occurrence of each item over the past two weeks using a five-point Likert scale (1 – None of the time, 2 – Rarely, 3 – Some of the time, 4 – Often, 5 – All of the time). The total score is obtained by summing the quotations for each item, with a higher total score indicating a higher level of psychological well-being [75]. The original British scale as well as the French translated version used in this research exhibited a single factor and good internal consistency (Cronbach’s alpha ranging from 0.85 to 0.89 across three samples for the French translation). In addition, the French WEMWBS proved to be sensitive in detecting changes following a mental health intervention in healthy and schizophrenia-remitted populations [76].
- **PANAS**: the Positive and Negative Affect Schedule contains two 10-item adjective subscales relating to different attractive and aversive moods that one can experience. Positive Affect (PA) reflects feelings of enthusiasm, activity, and alertness, while Negative Affect (NA) indicates distress and unpleasant engagement [77]. There has been debate over the scale’s structure, with some suggesting a two-factor model (PA and NA) and others proposing a three-factor model, which separates NA into “Afraid” and “Upset” dimensions [78–80]. For the French translation used in this research, the internal consistency coefficients were high: PA, Cronbach’s alpha = 0.90; 0.91 across two samples, NA – Global = 0.80; 0.84, NA – Upset = 0.81; 0.82, NA – Afraid = 0.76; 0.74 [80]. Participants were invited to express the extent to which they had experienced each of the 20 proposed moods in the past month using a five-point Likert scale: 1 - Very slightly or not at all, 2 - A little, 3 - Moderately, 4 - Quite a bit, 5 - Very much. The mean score of PA and NA factors was calculated.
- **WCS**: The Watts Connectedness Scale (WCS) is a psychometric tool developed from psychedelic research, particularly studies of psilocybin for treatment-resistant depression. Participants in psilocybin studies reported feeling an increased sense of “connectedness,” which has been identified as a key factor in their feeling of mental-health improvement. This sense of connectedness includes three aspects: connection to self, others, and the world [43,54]. The WCS includes 19 statements about emotional, community, and physical sensations, with participants rating their agreement on a 0-100 visual analogue scale. Scores are calculated for each of the three dimensions of connectedness, as well as an overall score [81,82]. The structure of the scale has been validated with high internal consistency. A 3-factor structure loading into a second-order WCS total score was found to be acceptable, while Cronbach’s alphas for the three first-order latent factors were as follows: 0.84 (Connectedness to Self), 0.87 (Connectedness to Others), 0.90 (Connectedness to the World). Additional composite reliabilities were 0.79 (Self), 0.87 (Others), 0.90 (World), 0.86 (second-order latent variable subsuming all three subscales, i.e. global connectedness) [82]. As a validated French version of the WCS does not yet exist, participants were presented with an in-house French translation of the WCS.
- **NR-6**: The 6-item Nature Relatedness (NR-6) scale is a shortened version of the original 21-item scale developed by E. Nisbet and colleagues [83]. The six items were chosen for their ability to differentiate between individuals with low and high nature-relatedness and their correlation with similar scales. These items cover two of the three dimensions in the original scale: “Self” (identification with nature) and “Experience” (contact with nature). Four items focus on self-identification with nature, while two address the need for nature and comfort with wilderness [84]. The NR-6 has shown good psychometric properties, with Cronbach’s alpha values ranging from 0.83 to 0.86 and strong correlations with the full 21-item scale [84]. Since a validated French version doesn’t exist, participants used an in-house translation and rated their agreement on a 5-point Likert scale (from 1 = strongly disagree, to 5 = strongly agree). The scores for each item were averaged to compute the participants’ NR-6 scores. Higher scores indicate a stronger connection with nature [84].

## Statistics

### Data analysis

We performed different types of general linear models (GLMs) depending on the nature of measures (i.e. continuous, binomial, multinomial distributions). We will detail each of these cases in the following paragraph. All these models were run with **Age, Gender, IPSE, Group** and their interactions defined as independent variables. Bayes Factors (BF) were used to compare and select the best models, applying Jeffreys’ classification with a decision threshold set at a BF ratio of 3:1—indicating ‘substantial’ evidence in favour of the preferred model over alternatives[85].

We tested two sets of models. In the first set, our main variable of interest, ***AICT training***, representing whether the participant received the training or not, was treated as binary. In the second set, the variable ***AICT Practice Duration*** was considered continuous, and represented the number of months practicing AICT since training. For both approaches, we conducted a model comparison using the WAIC index to determine which model provided the best fit to the data. For each analysis, model selection was made with Bayes Factor comparing a set of thirty linear models from the most complete model: *Test ∼ Gender +IPSE +Age\*****Duration****_AICT Practice_ +Practice_AutoHypnosis_ +Practice_Meditation_ +Practice_Relaxation_ +Exp_Trauma_ +Exp_LifeDanger_ +Exp_SpontaneousTrance_*, to the simplest: *Test* ∼ 1. We checked for the convergence of Markov Monte Carlo chains (MCMC). The extraction of data and plots were performed with Python3 [86] and statistical analysis were conducted with R (version 4.2.1) using the brms, multcomp, vegan and emmeans packages [87].

In both sets, Practice and Exp variables (*Practice_AutoHypnosis_ +Practice_Meditation_ +Practice_Relaxation_ +Exp_Trauma_ +Exp_LifeDanger_ +Exp_SpontaneousTrance)_*) were binary control variables. Inclusion of these control variables aimed at discriminating possible confounding influences carried by NOSCs or potential NOSC-inducing events apart from AICT on interest variables. Details about control variables included in statistical models are provided in Supplementary Materials.

### Bayesian Estimation and Inference Approach

To assess the existence and directionality of effects, we use the **probability of direction** (pd), which reflects the proportion of the posterior distribution that is strictly above or below zero. The pd can be interpreted as a Bayesian equivalent to a p-value: higher values (e.g., pd > 0.975) indicate greater certainty about the sign of the effect. While pd values are reported for all parameters, we focused **Bayes Factor (BF)** comparisons on our main parameter(s) of interest. For these, we compared the full model to a nested model excluding the relevant predictor. The resulting BFs quantify the relative evidence in favour of including the parameter (H₁) versus excluding it (H₀), with standard interpretive thresholds (e.g., 3 < BF < 10 = moderate evidence, BF > 10 = strong evidence). This hybrid strategy provides full transparency for all estimated effects while maintaining focused inference on key hypotheses.

### Ethic statement

The Research Ethics Committee of the University of Strasbourg approved the study protocol and registered the research project under the accreditation number Unistra/CER/2024-04. No access to the survey was allowed without the participant’s consent.

## Results

### Sociodemographic data

The online questionnaire was accessed 1004 times, allowing the analysis of 607 valid responses, 171 of which were from untrained participants, and 436 from AICT-trained participants (see **Table 1**). Most participants in the pool of valid responses reported being females (73.48%; n=446). The mean age of AICT-trained (48.99 ± 10.77 years old) and untrained participants (48.98 ± 10.69 years old) was similar, with a global mean of 48.99 ± 10.74 years of age. The mean individual socioeconomic level index (IPSE) in our sample (86.10 ± 13.77, AICT-trained participants: 86.45 ± 13.59, untrained participants: 85.19 ± 14.24) was above the cut-off value of 80, indicating a population of higher socioeconomic status [59]. Most participants reported practicing other NOSC, with more than half of them also practicing meditation (62.27%), 39.04% practicing relaxation methods, 22.08% practicing self-hypnosis, and 5.60% reporting the use of psychedelic substances. Reports of other NOSC practices were twice as-common among AICT-trained participants as among untrained participants. Examination of AICT practice frequency in AICT practitioners revealed that the proportion of individuals who had not practiced AICT for more than 6 months was negligible (n=6, data not shown), therefore informing that participants involved in this study were active AICT practitioners.

**Table 1.** Analysed sample descriptive sociodemographic data.

| Training<br>in AICT<br>↓ | Participants<br>n<br>(%) | Genders |  |  | Age<br>Mean ±<br>SD | Individual<br>socioeconomic<br>level index<br>Mean ± SD | Other NSC practices |  |  |  |
| --- | --- | --- | --- | --- | --- | --- | --- | --- | --- | --- |
|  |  | Women | Men | Non-<br>binary |  |  | Meditation | Self-Hypnosis | Relaxation,<br>Yoga, Tai-Chi | Psychedelic<br>substances<br>use |
|  |  | n<br>(%) | n<br>(%) | n<br>(%) |  |  | n<br>(%) | n<br>(%) | n<br>(%) | n<br>(%) |
| Not<br>trained | 171<br>(28.17) | 124<br>(27.80) | 45<br>(29.22) | 2<br>(28.57) | 48.98<br>±10.69 | 85.19<br>± 14.24 | 122<br>(20.10) | 37<br>(6.10) | 74<br>(12.19) | 13<br>(2.14) |
| Trained | 436<br>(71.83) | 322<br>(72.20) | 109<br>(70.78) | 5<br>(71.43) | 48.99<br>±10.77 | 86.45<br>± 13.59 | 256<br>(42.17) | 97<br>(15.98) | 163<br>(26.85) | 21<br>(3.46) |
| **Total** | 607<br>(100) | 446<br>(73.48) | 154<br>(25.37) | 7<br>(1.15) | 48.99<br>±10.74 | 86.10<br>± 13.77 | 378<br>(62.27) | 134<br>(22.08) | 237<br>(39.04) | 34<br>(5.60) |

### Evaluating the effect of the AICT training

For each winning Bayesian model, we examined the 95% credibility intervals (CrI) and the probability of direction (pd) to assess parameter significance. The results of this model comparison are presented in the Appendix, along with heatmap visualisations illustrating the differences in WAIC values between models. For each model, we report the *posterior median* and the *95% credible interval (CI)* for all fixed effects.

The results of the Bayesian models treating *AICT Training* as a binary variable are summarised in **Table 2** and illustrated by **figure 2**. Several parameters showed significant effects, as indicated by their credible intervals (CIs) not overlapping zero, probability of direction (pd) > 0.975 and Bayes Factor (BF) >3. Notably, ***AICT Training*** had a significant effect on ***RSE*** (0.91, 95% CrI [0.23, 1.6], pd=1.00, BF=9.3), ***WCS Self*** (5.2, 95% CrI [2.4, 8], pd=1.00, BF=1.8e2), ***WCS Others*** (6.2, 95% CrI [3, 9.3], pd=1.00, BF=5.1e2), ***WCS World*** (5.7, 95% CrI [2.1, 9.3], pd=1.00, BF=4.2e1), ***WCS Global*** (6.1, 95% CrI [3.4, 8.7], pd=1.00, BF=5.1e3) suggesting that, on average, AICT training increases RSE scores by 0.91 points, WCS Connection to Self-scores by 5.2 points, WCS Connection to Others by 6.2 points, WCS Connection to World by 5.7 points and WCS Global by 6.1 points.

**Figure 2.**
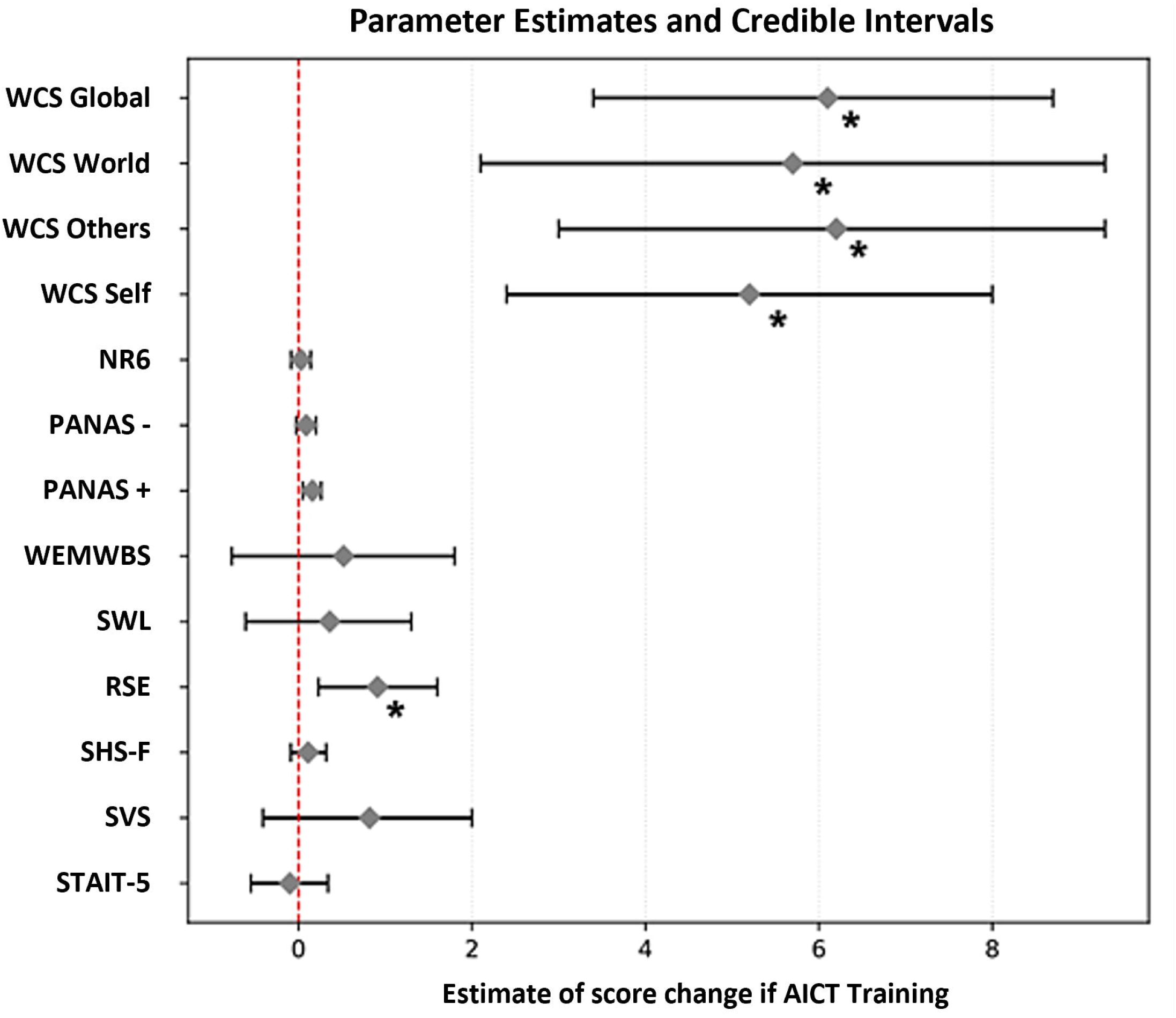
Posterior estimates and 95% credible intervals (CIs) for the effect of **AICT Training** across different models. Each point represents the posterior mean of the parameter estimate, and the horizontal bars indicate the 95% CIs. The dashed red vertical line at zero represents the null value (no effect). Estimates whose CIs do not overlap with zero suggest a more robust directional effect in the corresponding model and the ones with **Bayes Factors (BF) > 3**, indicating moderate to strong evidence for an effect, are marked with an asterisk (*).

**Table 2.**
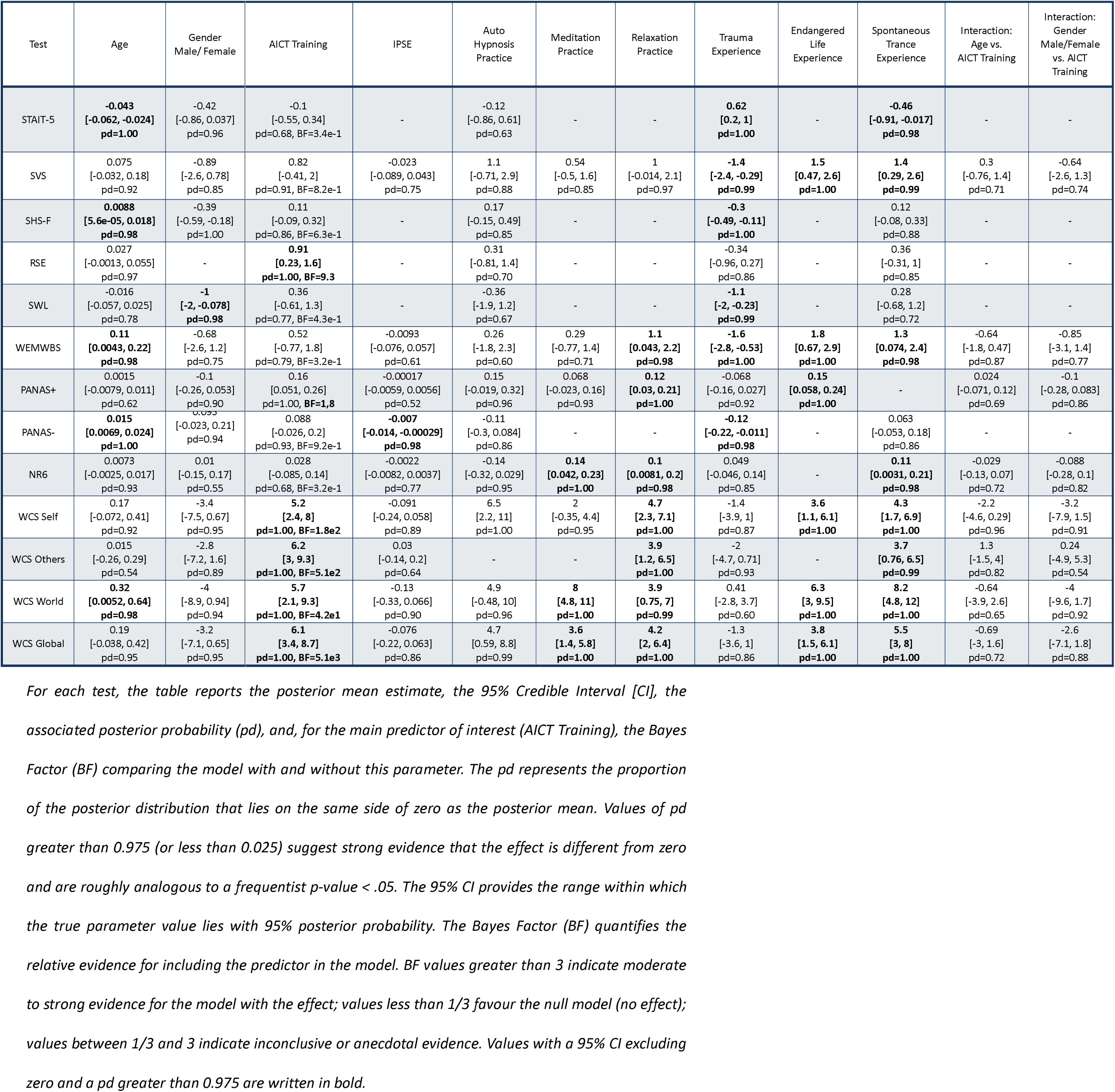
Summary of posterior estimates for each parameter across all models with AICT Training.

The indexes provided by CIs and pd also enabled the identification of demographic covariates that shared common effects with ***AICT Training*** on the investigated dimensions of well-being. Notably, ***Age*** had a significant effect on ***WCS World*** scores (0.32, 95% CI [0.0052, 0.64], pd=0.98), indicating that each additional year of age is associated with an increase of 0.32 points in ***WCS World*** scores. Despite the separate effects of ***AICT Training*** and ***Age*** on ***WCS World*** scores, no interaction effects were observed for this variable (***Age vs. AICT Training*** on ***WCS World,*** −0.64 [−3.9, 2.6] pd=0.65). Additionally, ***Gender*** showed no concurrent or interaction effects on variables influenced by ***AICT Training***. No concurrent effects on variables influenced by ***AICT Training*** were associated with ***IPSE***.

### Evaluating the effect of AICT Practice Duration

The results of the Bayesian models considering AICT Practice Duration as a continuous variable are summarised in **Table 3** and illustrated by **Figure 3**. The ***AICT Practice Duration*** parameter exhibited significant effects on several outcomes, with ***STAIT-5*** (−0.021, 95% CI [−0.037, −0.0045], pd=0.99, BF=3,2), ***RSE*** (0.046, 95% CI [0.022, 0.07] pd=1.00, BF=1,3e2), ***PANAS+*** (0.0054, 95% CI [0.0012, 0.0096] pd=0.99, BF=3.4), ***WCS Self*** (0.17, 95% CI [0.058, 0.28]) pd=1.00, BF=1.2e1), ***WCS Others*** (0.17, 95% CI [0.068, 0.28] pd = 1.00, BF=2,3e1), ***WCS World*** (0.22, 95% CI [0.07, 0.36] pd=1.00, BF=1.2e1), and ***WCS Global*** (0.2, 95% CI [0.095, 0.3] pd=1.00, BF=1.5e2). These results suggest that each additional month of practice is associated with a 0.021-point decrease in STAIT-5 scores and increases of 0.046 points in RSE, 0.0054 points in PANAS+, 0.17 points in WCS Self, 0.17 points in WCS Others, 0.22 points in WCS World and 0.2 points in *WCS Global*.

**Figure 3.**
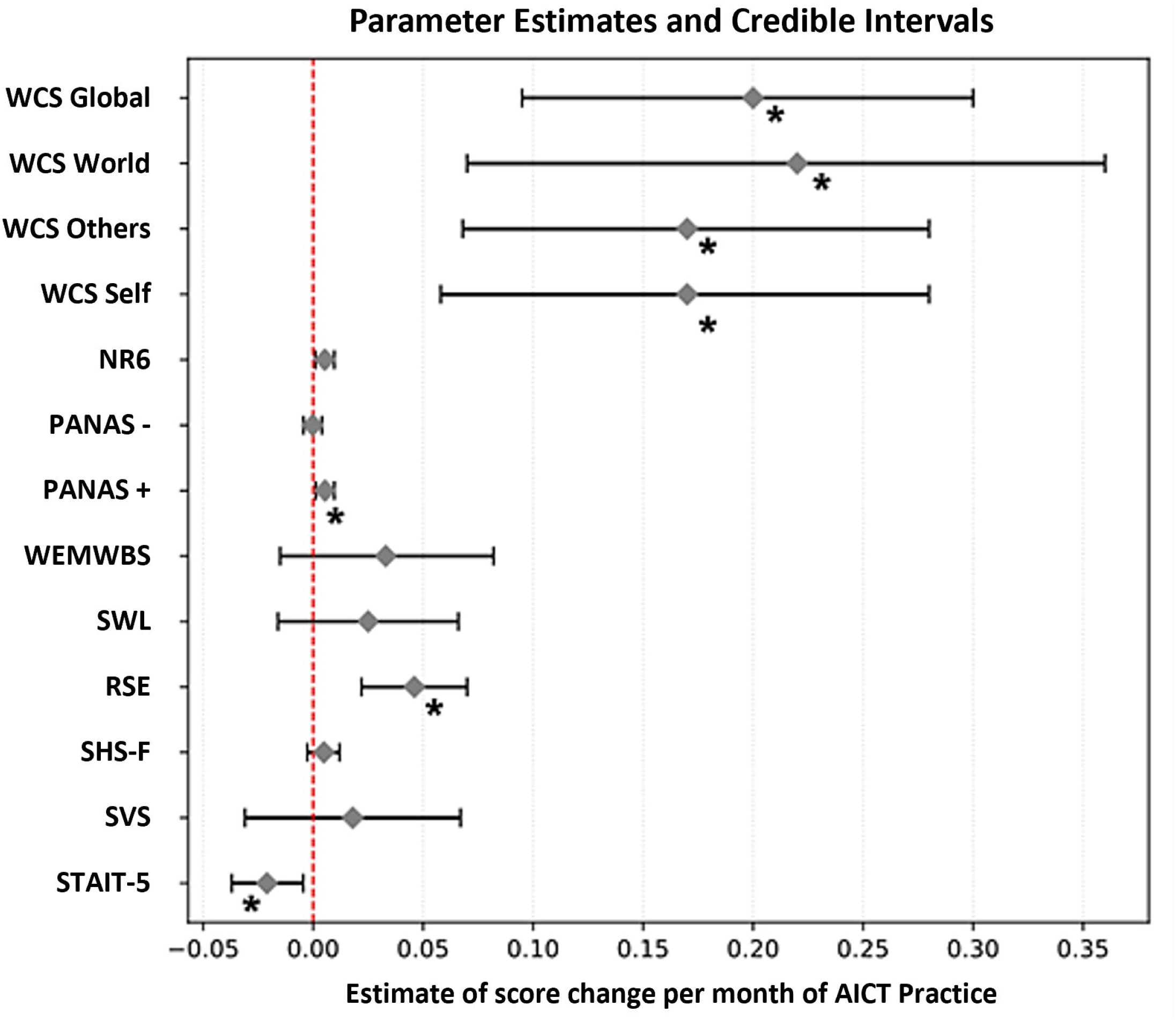
Posterior estimates and 95% credible intervals (CIs) for the effect of **AICT Practice Duration** across different models. Each point represents the posterior mean of the parameter estimate, and the horizontal bars indicate the 95% CIs. The dashed red vertical line at zero represents the null value (no effect). Estimates whose CIs do not overlap with zero suggest a more robust directional effect in the corresponding model and the ones with **Bayes Factors (BF) > 3**, indicating moderate to strong evidence for an effect, are marked with an asterisk (*).

**Table 3.**
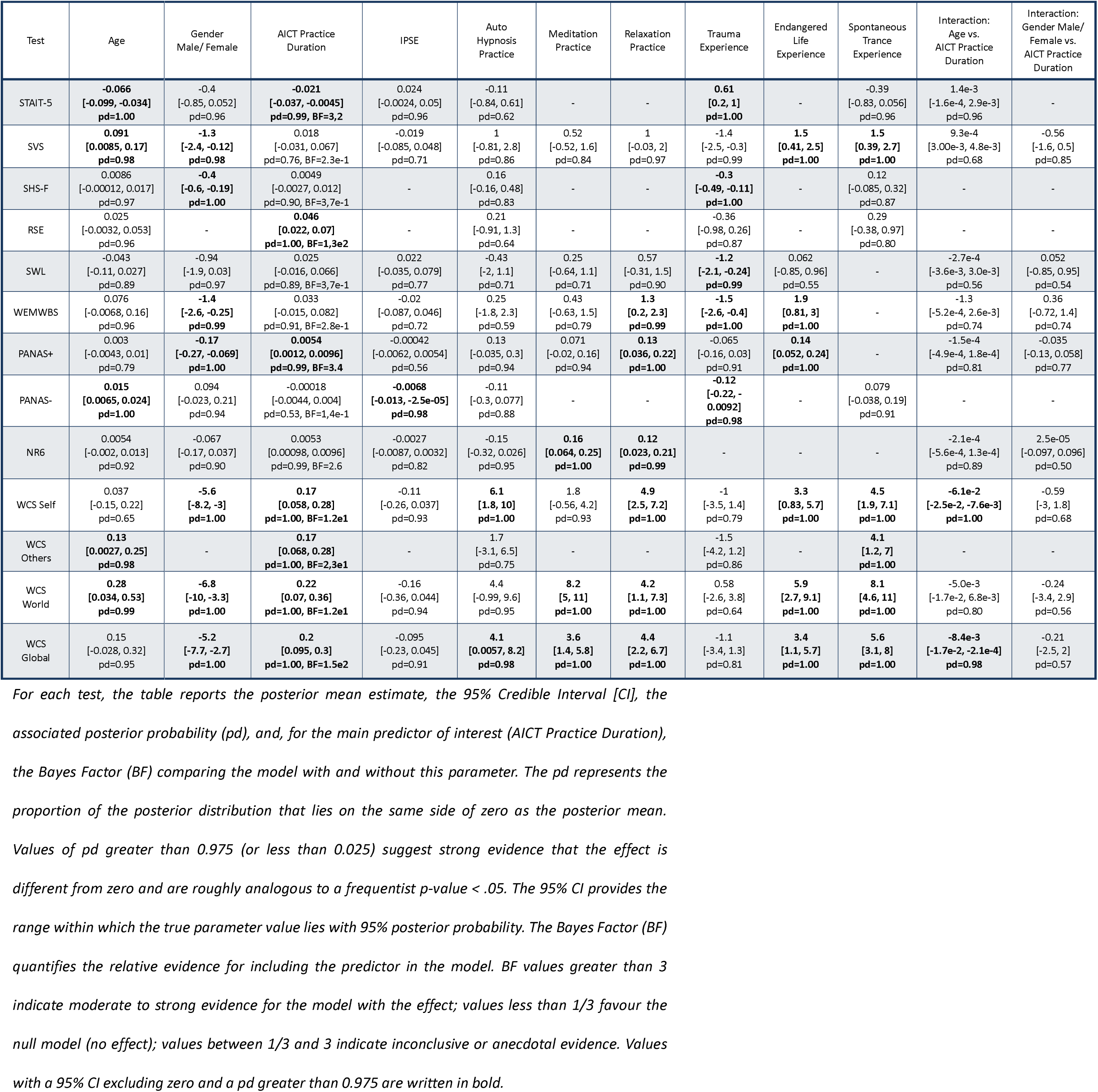
Summary of posterior estimates for each parameter across all models with AICT Practice Duration.

Additionally, the ***Age*** covariable showed effects on ***STAIT-5*** (−0.066, 95% CI [−0.099, −0.034] pd=1.00), ***WCS Others*** (0.13, 95% CI [0.0027, 0.25] pd=0.98), and ***WCS World*** (0.28, 95% CI [0.034, 0.53], pd=0.99), suggesting that each additional year of age is associated with a 0.066-point decrease in STAIT-5 scores and increases of 0.13 points in WCS Other, and 0.28 points in WCS World. The Interaction of ***Age*** and ***AICT Practice Duration*** (***Interaction: Age vs. AICT Practice Duration***) on ***WCS Self*** (−1.6e-2, 95% CI [−2.5e-2, −7.6e-3] pd=1.00) and ***WCS Global*** (−8.4e-3, 95% CI [−1.7e-2, −2.1e-4], pd=0.98) suggest that either each additional year of aging for a same AICT Practice Duration, or each additional year of AICT Practice Duration for a same age results in a 0.016-point decrease in ***WCS Self*** and a 0.0084-point decrease in ***WCS Global*** scores. However, the effects of ***Interaction: Age vs. AICT Practice Duration*** appear to be much weaker and only slightly counterbalance that of ***AICT Practice Duration***.

## Discussion

This research was conducted to evaluate whether the practice of a trance NOSC, AICT, may be related to a rise in the expression of subdimensions either part of the construct or contributing to a general state of well-being. The results offer a first quantitative description of well-being dimensions in AICT practitioners, comparatively to an age, gender, and socio-economically equivalent population of non-practitioners. The use of statistical modelling enabled the longitudinal analysis of variables related to well-being with respect to AICT practice duration, in addition to a general comparison between AICT-trained and untrained participants. The evaluation of AICT as a binary (*AICT Training*) or time-dependent (*AICT Practice Duration*) effector allowed for the identification of strong effects on connectedness (WCS) and self-esteem (RSE), to which more subtle effects on positive affects (PANAS+) and trait anxiety (STAIT-5) added-up when considering the influence of prolonged AICT practice.

Among the results unravelled, AICT Training appeared as associated with an increased general sense of connectedness, as well as increased connectedness across all subdimensions of the construct on trained participants (Self, Others, World). The constructs questioned by the WCS take root in the statements provided by depressed patients undergoing psychedelic research with psilocybin, which pointed to a particular breakthrough supported by an increased or renewed sense of connectedness. More precisely, the sense of connectedness described by patients refers to a global sense of body, mind and emotional awareness (connectedness to self), as well as feelings of understanding and inclusion within physical and social environments (connectedness to others, connectedness to world)[43,82]. Increased connectedness to self, world and others consequently to AICT practice may be explained by experiences lived during AICT-trance states in trained participants. One of the phenomena associated with AICT includes feelings of unity or bodily boundaries blurring/dissolution leading to a feeling of merging with the environment, firstly reported by the designer of the AICT method[88] and observed in subsequent research on AICT practitioners[33]. For instance, a sense of unity/harmony has been found to be common to NDE-experiencers and AICT practitioners[37], and a recent mixed-method study reported that AICT-trained participants experienced attenuated bodily boundaries and heightened feelings of oneness during the state of AICT comparatively to their ordinary state of consciousness[39]. Secondly, heightened feelings of connectedness to self might be explained by the coupling between changes on dimensions related to self-awareness (increased absorptive state, increased interoceptive and emotional awareness) and autonomic nervous system (ANS) activity changes reported in the AICT state[36], such that entranced individuals experience a state of immersion in self-centred stimuli. Being completely oneself/a sense of unicity has been reported by the majority (72%) of AICT trained participants of a recent group study[33] (see also[89]). Interestingly, while the state of increased bodily awareness and dissolution of body boundaries may seem paradoxical, they rather highlight a change in the nature of self-related perceptions than a loss of self-related perceptions in the AICT state (‘*my head was becoming waves*’)[33,35]. The already identified relationship between absorption and empathy[90] may therefore be potentiated by the absorptive experience of becoming another elicited by AICT, as supported by the increased feeling of empathy and openness to others/the world attributed by AICT practitioners [33]. Finally, experiences of AICT can also involve interactions with perceived presences, living beings or inanimate objects[33,35,91] which therefore confront practitioners with social situations involving a wider assembly not limited to human peers. The possibility of a dialogue with any existing thing is not without reminding the animistic vision of creation shared in shamanic cultures, which support the position of humankind as included in an interacting conscious whole[92]. While the relationship between interactive situations lived in trance states and the animistic interconnected vision of creation is beyond the scope of this research, it seems plausible that the state of connectedness with the world reported by AICT practitioners may stem – partially or fully – on widened experiences of interaction lived in AICT trance states. In support of this are similar reports of interactive experiences with disincarnated presences and increased connectedness described by patients undergoing psilocybin assisted therapy[43,55] and in psychedelic experiences induced by N,N-dimethyltryptamine (DMT)[93]. In addition, according to a survey study[94], psychedelic experiences induce belief changes in favour of the existence of non-physical conscious entities, a conscious universe, and consciousness in mammals and inanimate natural objects [94]. AICT and psychedelic-induced trances may then similarly foster connectedness to world by favouring experiences of reciprocal interaction between trance-experiencers and their surroundings. Interestingly, the similarity of outcomes on connectedness feelings during psychedelics and AICT states therefore raises the question of whether these NOSC may stimulate common neurophysiological systems.

Additionally, comparison of AICT Trained and Untrained participants highlighted increases in scores of self-esteem whenever AICT was considered as binary or time dependent. Similarly to connectedness, the consistence of this effect on both all-merged data irrespective of time, and time-dependent data, sustains the possibility of an influence of AICT practice readily accessible to most AICT practitioners regardless of practice duration. Such quick effects on self-esteem after a mind-body intervention have also been observed as soon as after a two-week yoga intervention[95] and after a 10-minutes short mindfulness meditation intervention[96]. The efficiency of a short 15-minutes mindfulness meditation intervention has also been challenged with conclusive results on dimensions of emotional processing[97]. Although the underlying mechanisms of AICT on self-esteem have not yet been elucidated, results provided by a chronic pain case-study suggest that AICT may simultaneously act as a buffer on self-related hindering processes and a booster by increasing empowerment. For instance, the patient involved in this study reported re-experiencing metaphorically his painful body, allowing for a reappraisal of his relationship with pain towards a more respectful rapport. He also mentioned the feeling that AICT helped him to soothe and overcome pain, and to accept more his fragilities[98]. The account of gaining control over environmental discomfort during AICT was also reported in a TMS-EEG case-report (“*there was a spiral that tried to catch the little woodpecker that was on my head (note: the TMS)*”[32]) and through a feeling of ‘repairing perceived dissonances’[33]. Furthermore, the phenomenology of AICT shows that this state also involves dealing with strong emotional and psychological content of pleasant or uncomfortable valences[33], in which movement and vocal productions – sounds, songs, speaking in tongues – provide a way of coping or interacting with the emerging matter[98]. The ability to cope may also be facilitated by the state of dissociation elicited by AICT, through which participants experience a dual viewpoint combining both embodied and witness experiences and dampened self-criticism[39]. This change of perspective is not without reminding the process of “decentring” occurring in meditative practices, known to elicit a process of cognitive defusion[99,100]. The relaxation of the outlines of the self experienced during AICT combined with cognitive defusion may then offer the opportunity to experience oneself as part of a whole and trigger humbling experiences susceptible to dedramatize errors and alleviate the importance given to self, as supported by evidence associated with AICT (“*I am everything and I am nothing, and I don’t have to worry about me and everything. It’s so big and so small, it’s comforting. I’m part of this world, no matter what form I take. We’re all part of it*”) [91]. Altogether, these reports support the practice of AICT as an enabling technique which positions the practitioner as a resourceful agent of their subjective experience. The state of capacity, autonomy and connectedness experienced through the state of AICT may then contribute to the satisfaction of the basic psychological needs of competence, autonomy and relatedness lying at the root of self-determination theory[101], which allows individuals to act from their integrated or autonomous sense of self, and build a sense of genuine self-esteem[102]. In providing a way to manage difficulties, AICT may also contribute to the improvement of emotional stability, which was shown to act as a mediator of well-being by favouring self-esteem[49]. This hypothesis is supported by reports from another research on AICT, which highlighted that participants expressed feeling changed towards being more serene, stable and self-confident[33], and by our current data displaying an increase of so-called “positive” affects (PANAS+) and a decrease of trait anxiety (STAIT-5) under the effect of the time of AICT practice.

Such time-dependent effects may reflect neurophysiological mechanisms resulting from repeated or sustained stimulation as observed with prolonged meditation practice[3,34,103]. Early effects of AICT may also result from the rather intensive training spanning on two-weeks-apart two sessions of two days, since strong effects lasting up to one month could be observed on dimensions of mental well-being following an intensive yoga and meditation retreat[104]. Continued practice following trainings may then favour the maintenance of immediate effects. One reason for the appearance of time-dependent AICT benefits may also reside on the repeated exposure to intense emotional and ANS arousal states[33,36,44,45]. As observed with psychedelics, experiences of AICT may confront individuals with intense emotional releases which can be felt as challenging or uncomfortable, while allowing for a process of management for their underlying causes and the acknowledgment of internal regulation resources. As cited above, participants express getting original understandings and reappraisals while in the state of AICT, or the opportunity to fight or act through metaphorical actions[31,32,44,91,98]. Thus, AICT practitioners engage on a management of their internal states on a regular basis through gestures and emotional releases. Furthermore, the state of AICT has also been recently associated with increased probability for mystical-type experiences including feelings of harmony and oneness[33,89]. In the psychedelic context, the occurrence of emotional breakthrough and mystical experiences were shown to predict increases in well-being in a subsample of 75 participants with low well-being baseline scores[105], while mystical experiences akin to oceanic boundlessness predicted decreases in scores of depression in treatment-resistant depressive patients[106]. Since the practice of AICT is facilitated by the absence of any pharmacological or material needs, practitioners may easily re-expose themselves to the AICT state and get the opportunity to draw on the benefits of mystical type and emotional release experiences repeatedly. Although limited to a single case-study, improvement of depression and anxiety scores could however be observed as soon as after the completion of the four-days AICT practice in a chronic pain patient, pointing to the possibility of immediate effects on these variables[98]. The absence of an immediate effect on anxiety in our large sample may be explained by the high variability, both interindividual and across AICT experiences, of both phenomenology and autonomic nervous system responses[36]. Then, the appearance of AICT effects on anxiety over time in our large sample may as well mirror a progressive decrease in interindividual variability of AICT effects as practitioners settle in their practice. From an ANS standpoint, the repetition of ANS activation elicited by AICT practice[36,45] may also enforce adaptive capacities of the ANS[107,108], thus promoting a better sense of adjustment through circumstances and consequent decreased vulnerability to anxiety triggers. Finally, increased positive affect over time of AICT practice may also be explained by changes in emotion regulation strategies towards acceptance and reappraisal rather than suppression[98]. Benefits of reappraisal strategies were shown to contribute to increased positive affects comparatively to repressive ones[109], while acceptance was described as pivotal in the recovery of treatment-resistant depressive patients after psilocybin-assisted therapy[43]. Overall, the practice of AICT may then offer a setting for emotional regulation favourable to the experience of positive affective states on the long run. However, further research will be necessary to comprehend long-term cognitive, psychological and physiological effects of AICT practice, as current hypotheses rely on a limited set of studies.

This survey study was the first attempt to identify the potential influence of AICT on well-being dimensions using a quantitative approach. Several limitations must be considered when interpreting the acquired data. Firstly, this research used a survey design that relied on self-evaluations by participants via a web-based platform (LimeSurvey). Although the inclusion and exclusion criteria were clearly disclosed and participants were required to express their endorsement, no further controls were in place to verify the accuracy of the participants’ statements. In that line, this study aimed to control for participants’ predisposition towards AICT by advertising the call for participants through channels targeted to current and future trainees (e.g. mailing lists) and by displaying it on the training organisation’s website. Despite these precautions, the call for participants may have reached individuals who were not interested in AICT, who may have completed the questionnaire without any further possibility of identification. Consequently, we cannot confirm that our sample population was exclusively composed of participants with a shared interest in AICT. Similarly, this study lacks the means to compare AICT-trained and AICT-untrained participants, both interested and uninterested in AICT. Furthermore, it would have been useful to ask participants about their beliefs regarding the potential influence of AICT on well-being, in order to assess whether the respondents comprised both those who found AICT to be efficient and those who did not. Our study also shows discrepancies in the number of participants according to their time of practice, with fewer participants having practised for more than three years (Supplementary materials). While this can be explained by the small number of trainees at this level of seniority, it would be interesting to investigate whether other factors may have influenced the reduced number of responses. Finally, this study used validated translations of standardised questionnaires where available and in-house translations of validated questionnaires where a validated translation was non-existent. Consequently, the validity and rigour of the in-house translations will require further validation.

## Conclusions

This study investigated the influence of AICT training and AICT practice duration on well-being dimensions using a survey design comprising validated, standardised scales addressed to AICT-trained and untrained participants. Our results showed that AICT training was associated with increased self-esteem and overall connectedness, as well as the subdimensions of connectedness to self, others and the world. The duration of AICT practice was found to be associated with an increase in overall connectedness, as well as the subdimensions of connectedness, self-esteem, and positive affect, and a decrease in trait anxiety. Further research is needed to clarify the underlying mechanisms set in motion by AICT and to confirm the current findings with a more diverse sample of participants.

## Data Availability

The cleaned raw dataset, dataset files for script analysis and R markdown script files are available at Open Science Framework via https://doi.org/10.17605/OSF.IO/UA4HJ.

## Acknowledgements

Amandine Fernandez gratefully thanks Laurianne Tournier-Pinloche for her valuable feedback and critical reading.

## Supporting information captions

**Supplementary figure 1: Descriptive histogram of the sample of respondents by gender and AICT practice group.** This histogram categorises participants according to the following criteria: AICT Training (yes/no), time elapsed since AICT training/duration of practice (months), gender.

**Supplementary Figure 2: Heatmap of WAIC differences between models for each test with a binary variable for AICT Training.** The value displayed is the difference between the WAIC of each model and that of the best model for a given test (Δ WAIC = WAICi - WAICmin). A lower value indicates a better model fit (red) and the best model fit is at 0.0.

**Supplementary figure 3. Heatmap of WAIC differences between models for each test with a continuous variable for AICT Practice Duration.** The value displayed is the difference between the WAIC of each model and that of the best model for a given test (Δ WAIC = WAICi - WAICmin). A lower value indicates a better model fit (red) and the best model fit is at 0.0.

**Supplementary figure 4. Prior predictive checks for the STAIT-5 model.** The top panel displays quantile bands (10–90%, 20–80%, 30–70%, and 40–60%) of simulated outcome values (STAIT-5) from the prior predictive distribution. The middle panel shows the distribution of means across simulated datasets, with vertical lines indicating the minimum, maximum, and median of the observed data (blue). The bottom panel presents the distribution of standard deviations in simulated datasets, with a green vertical line marking the observed standard deviation. Overall, the simulations suggest that the specified priors generate plausible values for the STAIT-5 outcome.

**Supplementary figure 5. Posterior predictive check for the STAIT-5 model.** The top panel shows the observed histogram (in black) overlaid with posterior predictive quantile ribbons. The middle panel displays the distribution of the simulated means, with vertical lines representing the median, min, and max of the observed data. The bottom panel shows the distribution of posterior simulated standard deviations, with a vertical line for the observed SD.

## References

1. Moujaes F, Rieser NM, Phillips C, de Matos NMP, Brügger M, Dürler P, et al. Comparing Neural Correlates of Consciousness: From Psychedelics to Hypnosis and Meditation. Biol Psychiatry Cogn Neurosci Neuroimaging. 2024;9: 533–543. doi:10.1016/j.bpsc.2023.07.003

2. Franco Corso SJ, O’Malley KY, Subaiya S, Mayall D, Dakwar E. The role of non-ordinary states of consciousness occasioned by mind-body practices in mental health illness. J Affect Disord. 2023;335: 166–176. doi:10.1016/j.jad.2023.04.116

3. Taylor VA, Daneault V, Grant J, Scavone G, Breton E, Roffe-vidal S, et al. Impact of meditation training on the default mode network during a restful state. Soc Cogn Affect Neurosci. 2013;8: 4–14. doi:10.1093/scan/nsr087

4. Jha K, Kumar P, Kumar Y, Ganashree CP, Tripathi CB, Shrikant BK, et al. The effects of Rajyoga mindfulness meditation training on heart rate variability in panic disorder: A randomized controlled trial. Indian J Psychiatry. 2025;67: 310–315. doi:10.4103/indianjpsychiatry.indianjpsychiatry_323_24

5. Huong XV. Examining the Scientific Foundation of Mindfulness and Alternative Approaches for Improving Mental Well-Being. The American Journal of Social Science and Education Innovations. 2023;05: 65–71. doi:10.37547/tajssei/volume05issue11-06

6. Dorjee D. Defining contemplative science: The metacognitive self-regulatory capacity of the mind, context of meditation practice and modes of existential awareness. Front Psychol. 2016;7: 1–15. doi:10.3389/fpsyg.2016.01788

7. Dor-Ziderman Y, Schweitzer Y, Nave O, Trautwein F-M, Fulder S, Lutz A, et al. Training the embodied self in its impermanence: meditators evidence neurophysiological markers of death acceptance. Neurosci Conscious. 2025;2025. doi:10.1093/nc/niaf002

8. Dahl CJ, Lutz A, Davidson RJ. Reconstructing and deconstructing the self: cognitive mechanisms in meditation practice. Trends Cogn Sci. 2015;19: 515–523. doi:10.1016/j.tics.2015.07.001

9. Otani A. Hypnosis and mindfulness: The twain finally meet. American Journal of Clinical Hypnosis. 2016;58: 383–398. doi:10.1080/00029157.2015.1085364

10. Peters LG. Shamanism: Phenomenology of a spiritual discipline. Journal of Transpersonal Psychology. 1989;21: 115–137.

11. Somer E, Saadon M. Stambali: Dissociative Possession and Trance in a Tunisian Healing Dance. Transcult Psychiatry. 2000;37: 580–600. doi:10.1177/136346150003700406

12. Winkelman M. Anthropology, Shamanism and Hallucinogens - Winkelman. In: Grob CS, Grigsby J, editors. Handbook of Medical Hallucinogens. New-York: The Guilford Press; 2021. pp. 46–67. Available: https://www.researchgate.net/publication/351287995_Anthropology_Shamanism_and_Hallucinogens_In_Handbook_of_Medical_Hallucinogens_Edited_by_Charles_S_Grob_and_Jim_Grigsby_NY_Guilford_Press_Pp_46-67

13. Huels ER, Kim H, Lee UC, Bel-Bahar T, Colmenero A V., Nelson A, et al. Neural Correlates of the Shamanic State of Consciousness. Front Hum Neurosci. 2021;15: 1–16. doi:10.3389/fnhum.2021.610466

14. Gosseries O, Marie N, Lafon Y, Bicego A, Grégoire C, Oswald V, et al. Exploration of trance states: phenomenology, brain correlates, and clinical applications. Curr Opin Behav Sci. 2024;58: 101400. doi:10.1016/j.cobeha.2024.101400

15. Peters Lg. trance, initiation, and psychotherapy in Tamang shamanism. Am Ethnol. 1982;9: 21–46. doi:10.1525/ae.1982.9.1.02a00020

16. Rogerson R. In the Voices of the Ancestors: Izangoma Trance Processes and Embodied Narratives Towards Decolonization Praxis. Thesis, York University, Toronto. 2017.

17. Turner TD. Music and Trance as Methods for Engaging with Suffering. Ethos. 2020;48: 74–92. doi:10.1111/etho.12265

18. Mu Q. Pulled by God: Sound and Altered State of Consciousness in the Hälqä-Sohbät Ritual of Uyghur Sufis. European Journal of Musicology. 2022;20: 173–195. doi:10.5450/ejm.20.1.2021.173

19. De Oliveira Pinto T. “Making Ritual Drama:” Dance, Music, and Representation in Brazilian “candomblé” and “umbanda.” The World of Music. 1991;33: 70–88. doi:www.jstor.org/stable/43562778

20. Pavlov S V., Korenyok V V., Reva N V., Tumyalis A V., Loktev K V., Aftanas LI. Effects of long-term meditation practice on attentional biases towards emotional faces: An eye-tracking study. Cogn Emot. 2015;29: 807–815. doi:10.1080/02699931.2014.945903

21. Moore A, Malinowski P. Meditation, mindfulness and cognitive flexibility. Conscious Cogn. 2009;18: 176–186. doi:10.1016/j.concog.2008.12.008

22. Zhang Q, Wang Z, Wang X, Liu L, Zhang J, Zhou R. The effects of different stages of mindfulness meditation training on emotion regulation. Front Hum Neurosci. 2019;13: 1–8. doi:10.3389/fnhum.2019.00208

23. Campbell TS, Labelle LE, Bacon SL, Faris P, Carlson LE. Impact of Mindfulness-Based Stress Reduction (MBSR) on attention, rumination and resting blood pressure in women with cancer: A waitlist-controlled study. J Behav Med. 2012;35: 262–271. doi:10.1007/s10865-011-9357-1

24. Hoge EA, Bui E, Marques L, Metcalf CA, Morris LK, Robinaugh DJ, et al. Randomized controlled trial of mindfulness meditation for generalized anxiety disorder: Effects on anxiety and stress reactivity. Journal of Clinical Psychiatry. 2013;74: 786–792. doi:10.4088/JCP.12m08083

25. Krygier JR, Heathers JAJ, Shahrestani S, Abbott M, Gross JJ, Kemp AH. Mindfulness meditation, well-being, and heart rate variability: A preliminary investigation into the impact of intensive vipassana meditation. International Journal of Psychophysiology. 2013;89: 305–313. doi:10.1016/j.ijpsycho.2013.06.017

26. Amihai I, Kozhevnikov M. The influence of buddhist meditation traditions on the autonomic system and attention. BioMed Research International. Hindawi Publishing Corporation; 2015. doi:10.1155/2015/731579

27. Moore A, Gruber T, Derose J, Malinowski P. Regular, brief mindfulness meditation practice improves electrophysiological markers of attentional control. Front Hum Neurosci. 2012;6: 1–15. doi:10.3389/fnhum.2012.00018

28. Lardone A, Liparoti M, Sorrentino P, Minino R, Polverino A, Lopez ET, et al. Topological changes of brain network during mindfulness meditation: An exploratory source level magnetoencephalographic study. AIMS Neurosci. 2022;9: 25–263. doi:10.3934/Neuroscience.2022013

29. Kumar G. P, Panda R, Sharma K, Adarsh A, Annen J, Martial C, et al. Changes in high-order interaction measures of synergy and redundancy during non-ordinary states of consciousness induced by meditation, hypnosis, and auto-induced cognitive trance. Neuroimage. 2024;293. doi:10.1016/j.neuroimage.2024.120623

30. Seligman R. Distress, Dissociation, and Embodied Experience: Reconsidering the Pathways to Mediumship and Mental Health. Ethos. 2005;33: 71–99. doi:10.1525/eth.2005.33.1.071

31. Flor-Henry P, Shapiro Y, Sombrun C. Brain changes during a shamanic trance: Altered modes of consciousness, hemispheric laterality, and systemic psychobiology. Cogent Psychol. 2017;4: 1–25. doi:10.1080/23311908.2017.1313522

32. Gosseries O, Fecchio M, Wolff A, Sanz LRD, Sombrun C, Vanhaudenhuyse A, et al. Behavioural and brain responses in cognitive trance: A TMS-EEG case study. Clinical Neurophysiology. 2020;131: 586–588. doi:10.1016/j.clinph.2019.11.011

33. Gregoire C, Sombrun C, Lenaif P, Marie N, Giovine A, Walter M, et al. Phenomenological characteristics of auto-induced cognitive trance and Mahorikatan®trance. Neurosci Conscious. 2024;2024. doi:10.1093/nc/niae024

34. Kral TRA, Schuyler BS, Mumford JA, Rosenkranz MA, Lutz A, Davidson RJ. Impact of short- and long-term mindfulness meditation training on amygdala reactivity to emotional stimuli. Neuroimage. 2018;181: 301–313. doi:10.1016/j.neuroimage.2018.07.013

35. Vanhaudenhuyse A, Castillo M, Martial C, Annen J, Bicego A, Rousseaux F, et al. Phenomenology of auto-induced cognitive trance using text mining: a prospective and exploratory group study. Neurosci Conscious. 2024;2024. doi:10.1093/nc/niae036

36. Oswald V, Jerbi K, Sombrun C, Jitka A, Martial C, Gosseries O, et al. Understanding individual differences in non-ordinary state of consciousnesse: Relationship between phenomenological experiences and autonomic nervous system. International Journal of Clinical and Health Psychology. 2025;25: 100552. doi:10.1016/j.ijchp.2025.100552

37. Martial C, Simon J, Puttaert N, Gosseries O, Charland-Verville V, Nyssen AS, et al. The Near-Death Experience Content (NDE-C) scale: Development and psychometric validation. Conscious Cogn. 2020;86: 1–25. doi:10.1016/j.concog.2020.103049

38. Vanhaudenhuyse A, Castillo M-C, Martial C, Annen J, Bicego A, Rousseaux F, et al. Phenomenology of auto-induced cognitive trance using text mining: a prospective and exploratory group study - Supplementary materials. Neurosci Conscious. 2024;2024. doi:10.1093/nc/niae036

39. De Oliveira P, Meunier H, Breton A, Dambrun M. From the Attenuation of Self-Boundaries to Inner Peace: How Non-Ordinary States of Consciousness Modulates Self-experience and Subjective Well-Being. Submitted.

40. Dambrun M. When the dissolution of perceived body boundaries elicits happiness: The effect of selflessness induced by a body scan meditation. Conscious Cogn. 2016;46: 89–98. doi:10.1016/j.concog.2016.09.013

41. Mason NL, Kuypers KPC, Müller F, Reckweg J, Tse DHY, Toennes SW, et al. Me, myself, bye: regional alterations in glutamate and the experience of ego dissolution with psilocybin. Neuropsychopharmacology. 2020;45: 2003–2011. doi:10.1038/s41386-020-0718-8

42. Timmermann C, Roseman L, Williams L, Erritzoe D, Martial C, Cassol H, et al. DMT models the near-death experience. Front Psychol. 2018;9: 1–12. doi:10.3389/fpsyg.2018.01424

43. Watts R, Day C, Krzanowski J, Nutt D, Carhart-Harris R. Patients’ Accounts of Increased “Connectedness” and “Acceptance” After Psilocybin for Treatment-Resistant Depression. J Humanist Psychol. 2017;57: 520–564. doi:10.1177/0022167817709585

44. Baldaccini S. L’influence de la Transe cognitive auto-induite sur le bien-être et la relation thérapeutiquee: une étude préliminaire auprès de six soignants. Hegel. 2025;n° 154: 27–41. doi:10.3917/heg.154.0027

45. Oswald V, Vanhaudenhuyse A, Annen J, Martial C, Bicego A, Rousseaux F, et al. Autonomic nervous system modulation during self-induced non-ordinary states of consciousness. Sci Rep. 2023;13: 15811. doi:10.1038/s41598-023-42393-7

46. Kirmayer LJ. Mindfulness in cultural context. Transcult Psychiatry. 2015;52: 447–469. doi:10.1177/1363461515598949

47. Salvado M, Marques DL, Pires IM, Silva NM. Mindfulness-based interventions to reduce burnout in primary healthcare professionals: A systematic review and meta-analysis. Healthcare (Switzerland). 2021;9. doi:10.3390/healthcare9101342

48. Keune PM, Perczel Forintos D. Mindfulness meditation: A preliminary study on meditation practice during everyday life activities and its association with well-being. Psihologijske Teme. 2010;19: 373–386.

49. Bajaj B, Gupta R, Sengupta S. Emotional Stability and Self-Esteem as Mediators Between Mindfulness and Happiness. J Happiness Stud. 2019;20: 2211–2226. doi:10.1007/s10902-018-0046-4

50. Nourollahimoghadam E, Gorji S, Gorji A, Khaleghi Ghadiri M. Therapeutic role of yoga in neuropsychological disorders. World J Psychiatry. 2021;11: 754–773. doi:10.5498/wjp.v11.i10.754

51. Varambally S, Gangadhar BN. Yoga: A spiritual practice with therapeutic value in psychiatry. Asian J Psychiatr. 2012;5: 186–189. doi:10.1016/j.ajp.2012.05.003

52. Davis AK, Barrett FS, May DG, Cosimano MP, Sepeda ND, Johnson MW, et al. Effects of Psilocybin-Assisted Therapy on Major Depressive Disorder: A Randomized Clinical Trial. JAMA Psychiatry. 2021;78: 481–489. doi:10.1001/jamapsychiatry.2020.3285

53. Goodwin GM, Aaronson ST, Alvarez O, Arden PC, Baker A, Bennett JC, et al. Single-Dose Psilocybin for a Treatment-Resistant Episode of Major Depression. N Engl J Med. 2022;387: 1637–1648. doi:10.1056/NEJMoa2206443

54. Carhart-Harris RL, Erritzoe D, Haijen E, Kaelen M, Watts R. Psychedelics and connectedness. Psychopharmacology (Berl). 2018;235: 547–550. doi:10.1007/s00213-017-4701-y

55. Belser AB, Agin-Liebes G, Swift TC, Terrana S, Devenot N, Friedman HL, et al. Patient Experiences of Psilocybin-Assisted Psychotherapy: An Interpretative Phenomenological Analysis. J Humanist Psychol. 2017;57: 354–388. doi:10.1177/0022167817706884

56. Leamy M, Bird V, Le Boutillier C, Williams J, Slade M. Conceptual framework for personal recovery in mental health: Systematic review and narrative synthesis. British Journal of Psychiatry. 2011;199: 445–452. doi:10.1192/bjp.bp.110.083733

57. Lee S-H, Baldina E, Lee E, Youm Y. Social connectedness and hair cortisol in community-dwelling older adults. Compr Psychoneuroendocrinol. 2021;6: 100053. doi:10.1016/j.cpnec.2021.100053

58. Cervinka R, Röderer K, Hefler E. Are nature lovers happy? on various indicators of well-being and connectedness with nature. J Health Psychol. 2012;17: 379–388. doi:10.1177/1359105311416873

59. Genoud PA. Indice de position socioéconomiquee: un calcul simplifié [Socioeconomic status: a simplified calculation]. [En ligne]✉: www.unifr.ch/cerf/ipse. 2011; 1–9. Available: http://www.unifr.ch/ipg/assets/files/DocGenoud/IPSE.pdf

60. Harrell Fe, Lee Kl, Mark Db. Multivariable prognostic models: issues in developing models, evaluating assumptions and adequacy, and measuring and reducing errors. Stat Med. 1996;15: 361–387. doi:10.1002/(SICI)1097-0258(19960229)15:4<361::AID-SIM168>3.0.CO;2-4

61. Peduzzi P, Concato J, Kemper E, Holford TR, Feinstein AR. A simulation study of the number of events per variable in logistic regression analysis. J Clin Epidemiol. 1996;49: 1373–1379. doi:10.1016/S0895-4356(96)00236-3

62. Zsido AN, Teleki SA, Csokasi K, Rozsa S, Bandi SA. Development of the short version of the spielberger state—trait anxiety inventory. Psychiatry Res. 2020;291: 113223. doi:10.1016/j.psychres.2020.113223

63. Bruchon-Schweitzer M, Paulhan I. Inventaire d’anxiété état-trait forme Y (STAI-Y)e: [manuel]. Editions du Centre de psychologie Appliquée, editor. Paris; 1993. Available: https://www.sudoc.abes.fr/cbs/DB=2.1//SRCH?IKT=12&TRM=03584650X&COOKIE=U10178,Klecteurweb,D2.1,E1393e613-4e,I250,B341720009+,SY,QDEF,A%5C9008+1,,J,H2-26,,29,,34,,39,,44,,49-50,,53-78,,80-87,NLECTEUR+PSI,R10.34.103.180,FN

64. Uno T, Matsuo T, Asano M, Loh PY. Effects of Simulated Visual Impairment Conditions on Movement and Anxiety during Gap Crossing. Healthcare (Switzerland). 2024;12: 1–11. doi:10.3390/healthcare12010042

65. Kotsou I, Leys C. “Echelle de bonheur subjectif (SHS): Propriétés psychométriques de la version française de l’échelle (SHS-F) et ses relations avec le bien-être psychologique, l’affect et la dépression”. Can J Behav Sci. 2017;49: 1–6. doi:10.1037/cbs0000060

66. Lyubomirsky Sonja, Heidi L. A Measure of Subjective Happiness: Preliminary Reliability and Construct Validation. Soc Indic Res. 1999;46: 137–155.

67. Ryan RM, Frederick C. On Energy, Personality, and Health: Subjective Vitality as a Dynamic Reflection of Well-Being. J Pers. 1997;65: 529–565. doi:10.1111/j.1467-6494.1997.tb00326.x

68. Bostic TJ, Mc Gartland Rubio D, Hood M. A Validation of the Subjective Vitality Scale Using Structural Equation Modeling. Soc Indic Res. 2000;52: 313–324. doi:10.1023/A:1007136110218

69. Salama-younes M, Montazeri A, Ismaïl A, Roncin C. Factor structure and internal consistency of the 12-item General Health Questionnaire (GHQ-12) and the Subjective Vitality Scale ( VS), and the relationship between theme: a study from France. 2009;6: 1–7. doi:10.1186/1477-7525-7-22

70. Donnellan MB, Trzesniewski KH, Robins RW. Measures of Self-Esteem. Measures of Personality and Social Psychological Constructs. Elsevier Inc.; 2015. doi:10.1016/B978-0-12-386915-9.00006-1

71. Monteiro RP, de Coelho GL H, Hanel PHP, de Medeiros ED, da Silva PDG. The Efficient Assessment of Self-Esteem: Proposing the Brief Rosenberg Self-Esteem Scale. Appl Res Qual Life. 2022;17: 931–947. doi:10.1007/s11482-021-09936-4

72. Vallieres EF, Vallerand RJ. Traduction et Validation Canadienne-Française de L’échelle de L’estime de Soi de Rosenberg. International Journal of Psychology. 1990;25: 305–316. doi:10.1080/00207599008247865

73. Diener E, Emmons RA, Larsen RJ, Griffin S. The Satisfaction With Life Scale. J Pers Assess. 1985;49: 71–75. doi:10.1207/s15327752jpa4901_13

74. Blais MR, Vallerand RJ, Pelletier LG, Brière NM. L’échelle de satisfaction de vie: Validation canadienne-française du “Satisfaction with Life Scale.” Can J Behav Sci. 1989;21: 210–223. doi:10.1037/h0079854

75. Tennant R, Hiller L, Fishwick R, Platt S, Joseph S, Weich S, et al. The Warwick-Edinburgh mental well-being scale (WEMWBS): Development and UK validation. Health Qual Life Outcomes. 2007;5: 1–13. doi:10.1186/1477-7525-5-63

76. Trousselard M, Steiler D, Dutheil F, Claverie D, Canini F, Fenouillet F, et al. Validation of the Warwick-Edinburgh Mental Well-Being Scale (WEMWBS) in French psychiatric and general populations. Psychiatry Res. 2016;245: 282–290. doi:10.1016/j.psychres.2016.08.050

77. Watson D, Clark LA, Tellegen A. Development and validation of brief measures of positive and negative affect: The PANAS scales. J Pers Soc Psychol. 1988;54: 1063–1070. doi:10.1037/0022-3514.54.6.1063

78. Mehrabian A. Comparison of the PAD and PANAS as models for describing emotions and for differentiating anxiety from depression. J Psychopathol Behav Assess. 1997;19: 331–357. doi:10.1007/BF02229025

79. Killgore WDS. Evidence for a Third Factor on the Positive and Negative Affect Schedule in a College Student Sample. Percept Mot Skills. 2000;90: 147–152. doi:10.2466/pms.2000.90.1.147

80. Gaudreau P, Sanchez X, Blondin JP. Positive and negative affective states in a performance-related setting - Testing the factorial structure of the PANAS across two samples of French-Canadian participants. European Journal of Psychological Assessment. 2006;22: 240–249. doi:10.1027/1015-5759.22.4.240

81. Watts R. Watts connectedness scale. DrrosalindwattsCom. 2024; 2–4. Available: https://www.drrosalindwatts.com/watts-connectedness-scale

82. Watts R, Kettner H, Geerts D, Gandy S, Kartner L, Mertens L, et al. The Watts Connectedness Scale: a new scale for measuring a sense of connectedness to self, others, and world. Psychopharmacology (Berl). 2022. doi:10.1007/s00213-022-06187-5

83. Nisbet EK, Zelenski JM, Murphy SA. The nature relatedness scale: Linking individuals’ connection with nature to environmental concern and behavior. Environ Behav. 2009;41: 715– 740. doi:10.1177/0013916508318748

84. Nisbet EK, Zelenski JM. The NR-6: A new brief measure of nature relatedness. Front Psychol. 2013;4: 1–11. doi:10.3389/fpsyg.2013.00813

85. Andraszewicz S, Scheibehenne B, Rieskamp J, Grasman R, Verhagen J, Wagenmakers EJ. An Introduction to Bayesian Hypothesis Testing for Management Research. J Manage. 2015;41: 521–543. doi:10.1177/0149206314560412

86. Van Rossum G, Drake FL. Python 3 Reference Manual. CreateSpace Independent Publishing Platform; 2009. Available: https://books.google.fr/books/about/Python_3_Reference_Manual.html?id=KIybQQAACAAJ&redir_esc=y

87. R Core Team. R: A Language and Environment for Statistical Computing. Vienna, Austria: R Foundation for Statistical Computing; 2018. Available: https://www.r-project.org/

88. Sombrun C. La Diagonale De La Joie. Albin Michel; 2021. Available: https://www.fnac.com/a15275328/Corine-Sombrun-La-Diagonale-de-la-joie

89. Bicego A, Alnagger N, Cardeña E, Sombrun C, Annen J, Gosseries O, et al. Exploring Mystical-Type Experiences Through Auto-Induced Cognitive Trance. International Journal of Clinical and Experimental Hypnosis. 2025;00: 1–16. doi:10.1080/00207144.2025.2544055

90. Wickramasekera IE. Empathic features of absorption and incongruence. American Journal of Clinical Hypnosis. 2007;50: 59–69. doi:10.1080/00029157.2007.10401598

91. Ollagnier-Beldame M, Sombrun C. Revitalizing the Fleshly Bond to the Web of Life with Auto-Induced Cognitive Trance. World Futures. 2026; 1–23. doi:10.1080/02604027.2026.2622292

92. Harvey G. Animism rather than Shamanisme: New Approaches to what Shamans do (for other animists). First. In: Continuum, editor. Spirit Possession and Trancee: New Interdisciplinary Perspectives. First. London: Continuum; 2010. pp. 16–34. doi:10.5040/9781472549365.ch-002

93. Davis AK, Clifton JM, Weaver EG, Hurwitz ES, Johnson MW, Griffiths RR. Survey of entity encounter experiences occasioned by inhaled N,N-dimethyltryptamine: Phenomenology, interpretation, and enduring effects. Journal of Psychopharmacology. 2020;34: 1008–1020. doi:10.1177/0269881120916143

94. Nayak SM, Singh M, Yaden DB, Griffiths RR. Belief changes associated with psychedelic use. Journal of Psychopharmacology. 2023;37: 80–92. doi:10.1177/02698811221131989

95. Tejvani R, Metri K, Agrawal J, Nagendra H. Effect of Yoga on anxiety, depression and self-esteem in orphanage residents: A pilot study. AYU (An international quarterly journal of research in Ayurveda). 2016;37: 22. doi:10.4103/ayu.ayu_158_15

96. Hooper R, Guest E, Ramsey-Wade C, Slater A. A brief mindfulness meditation can ameliorate the effects of exposure to idealised social media images on self-esteem, mood, and body appreciation in young women: An online randomised controlled experiment. Body Image. 2024;49: 101702. doi:10.1016/j.bodyim.2024.101702

97. Wu R, Liu LL, Zhu H, Su WJ, Cao ZY, Zhong SY, et al. Brief Mindfulness Meditation Improves Emotion Processing. Front Neurosci. 2019;13: 1–10. doi:10.3389/fnins.2019.01074

98. Collignon G, Bicego A, Faymonville M, Gosseries O, Bonhomme V, Dive D, et al. Therapeutic Use of Auto-Induced Cognitive Trance in a Chronic Pain Setting: A Case Study Using Mixed Methodology. OBM Integrative and Complementary Medicine. 2025;010: 1–20. doi:10.21926/obm.icm.2501012

99. Timmermann C, Bauer PR, Gosseries O, Vanhaudenhuyse A, Vollenweider F, Laureys S, et al. A neurophenomenological approach to non-ordinary states of consciousness: hypnosis, meditation, and psychedelics. Trends Cogn Sci. 2023;27: 139–159. doi:10.1016/j.tics.2022.11.006

100. Forman EM, Herbert JD, Juarascio AS, Yeomans PD, Zebell JA, Goetter EM, et al. The Drexel defusion scale: A new measure of experiential distancing. J Contextual Behav Sci. 2012;1: 55–65. doi:10.1016/j.jcbs.2012.09.001

101. Deci EL, Ryan RM. The “What” and “Why” of Goal Pursuits: Human Needs and the Self-Determination of Behavior. 2000.

102. Deci EL, Ryan RM. Human autonomy: The basis for true self-esteem. In: Kernis MH, editor. Efficacy, agency, and self-esteem. New-York: Plenum Press; 1995. pp. 31–50.

103. Ehmann S, Sezer I, Treves IN, Gabrieli JDE, Sacchet MD. Mindfulness, cognition, and long-term meditators: Toward a science of advanced meditation. Imaging Neuroscience. 2025;3. doi:10.1162/IMAG.a.82

104. Sadhasivam S, Alankar S, Maturi R, Vishnubhotla R V., Mudigonda M, Pawale D, et al. Inner Engineering Practices and Advanced 4-day Isha Yoga Retreat Are Associated with Cannabimimetic Effects with Increased Endocannabinoids and Short-Term and Sustained Improvement in Mental Health: A Prospective Observational Study of Meditators. Evidence-based Complementary and Alternative Medicine. 2020;2020. doi:10.1155/2020/8438272

105. Roseman L, Haijen E, Idialu-Ikato K, Kaelen M, Watts R, Carhart-Harris R. Emotional breakthrough and psychedelics: Validation of the Emotional Breakthrough Inventory. Journal of Psychopharmacology. 2019;33: 1076–1087. doi:10.1177/0269881119855974

106. Roseman L, Nutt DJ, Carhart-Harris RL. Quality of acute psychedelic experience predicts therapeutic efficacy of psilocybin for treatment-resistant depression. Front Pharmacol. 2018;8. doi:10.3389/fphar.2017.00974

107. Décarie-Spain L, Hayes AMR, Lauer LT, Kanoski SE. The gut-brain axis and cognitive control: A role for the vagus nerve. Semin Cell Dev Biol. 2024;156: 201–209. doi:10.1016/j.semcdb.2023.02.004

108. Blase K, Vermetten E, Lehrer P, Gevirtz R. Neurophysiological approach by self-control of your stress-related autonomic nervous system with depression, stress and anxiety patients. Int J Environ Res Public Health. 2021;18. doi:10.3390/ijerph18073329

109. Gross JJ, John OP. Individual Differences in Two Emotion Regulation Processes: Implications for Affect, Relationships, and Well-Being. J Pers Soc Psychol. 2003;85: 348–362. doi:10.1037/0022-3514.85.2.348

