## Supporting information for "Trance practice and well-being measures: the case of Auto-Induced Cognitive Trance"

Files available on OSF (<https://doi.org/10.17605/OSF.IO/UA4HJ>):

Scripts for analyses - R Markdown files:

1) Analysis_Formation_TCAI_binary_final1.Rmd

2) Analysis_Formation_TCAI_final1.Rmd

Data files analysed through the abovementioned scripts:

3) Data_Treated.csv

4) Test_WCS.csv

Cleaned raw data of all valid responses, collected through LimeSurvey (from february to may 2024):

5) Cleaned_raw_dataset.xlsx

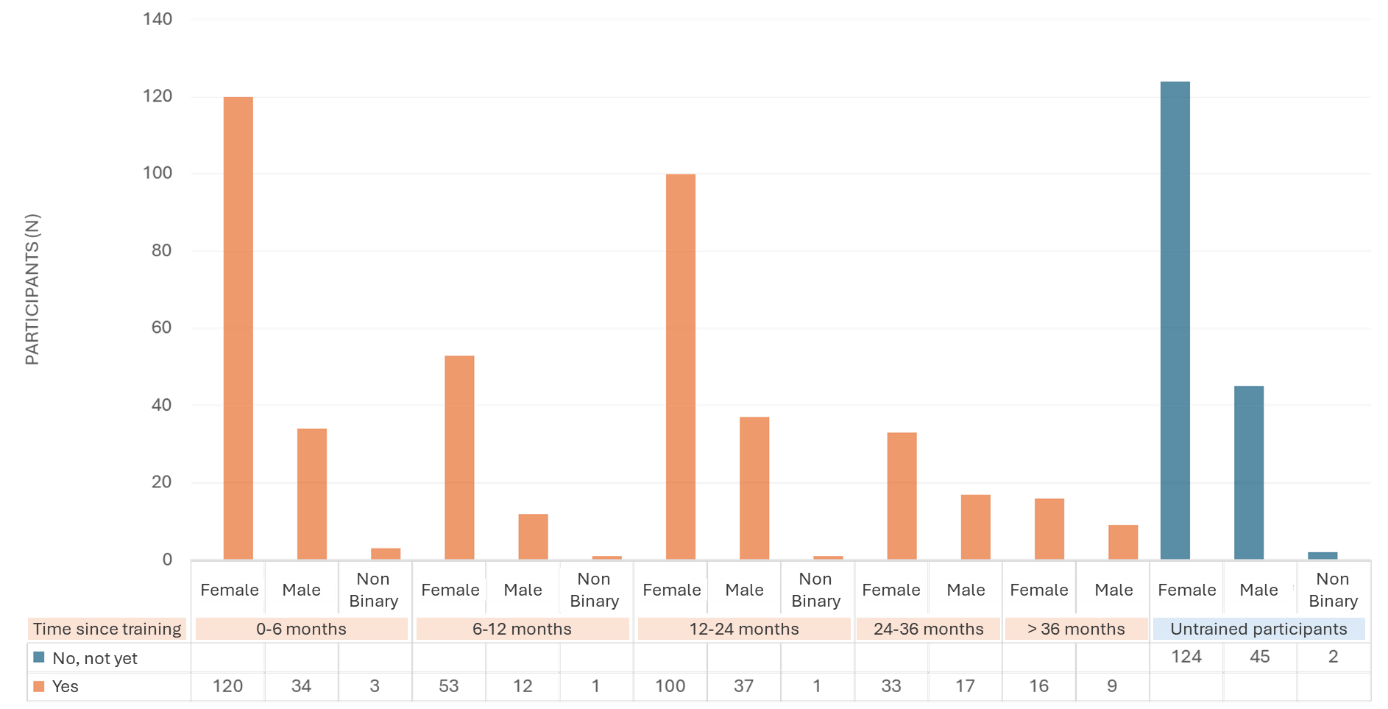

**Supplementary figure 1:** **Descriptive histogram of the sample of respondents by gender and AICT practice group.** This histogram categorises participants according to the following criteria: AICT Training (yes/no), time elapsed since AICT training/duration of practice (months), gender (Male/Female/Non Binary).

Model Selection using WAIC

To compare models and select the most appropriate one for each dependent variable, we relied on the **Watanabe-Akaike Information Criterion (WAIC)**, a fully Bayesian criterion for model comparison. WAIC estimates the out-of-sample predictive accuracy by penalising model complexity based on the effective number of parameters. Lower WAIC values indicate better expected predictive performance. Importantly, unlike classical information criteria (e.g., AIC), WAIC accounts for the full posterior distribution, making it suitable for evaluating hierarchical and non-linear Bayesian models. In each case, the model with the lowest WAIC was selected as the "winning" model for subsequent interpretation.

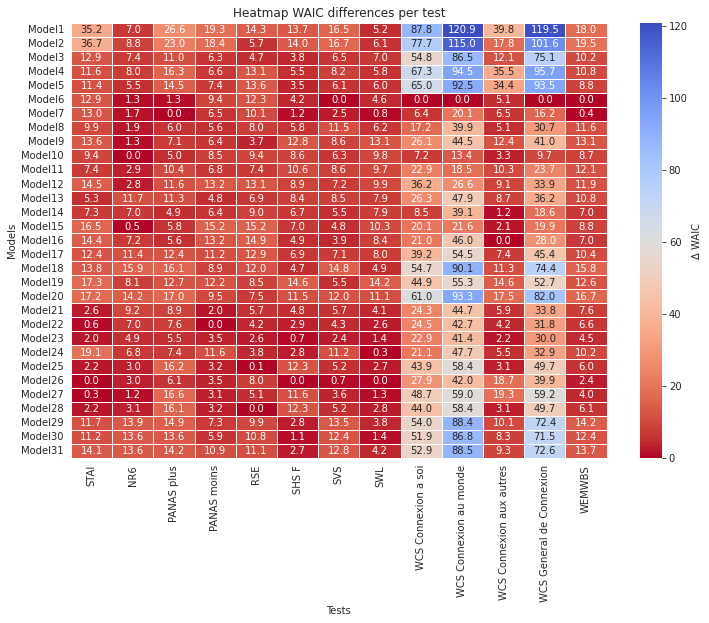
**Supplementary Figure 2: Heatmap of WAIC differences between models for each test with a binary variable for AICT Training.** The value displayed is the difference between the WAIC of each model and that of the best model for a given test (Δ WAIC = WAICi - WAICmin). A lower value indicates a better model fit (red) and the best model fit is at 0.0.

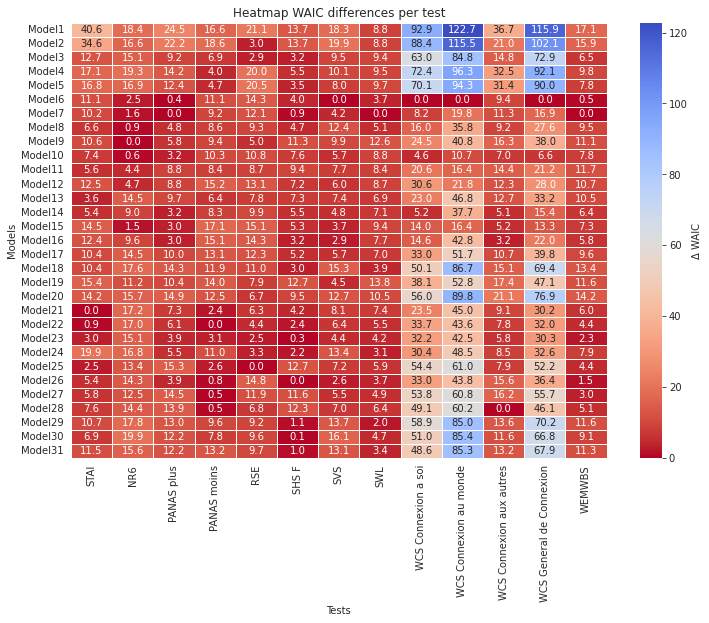
**Supplementary figure 3. Heatmap of WAIC differences between models for each test with a continuous variable for AICT Practice Duration.** The value displayed is the difference between the WAIC of each model and that of the best model for a given test (Δ WAIC = WAICi - WAICmin). A lower value indicates a better model fit (red) and the best model fit is at 0.0.

### Bayes factor workflow

Inspired from Schad, Daniel J, Michael Betancourt, and Shravan Vasishth. 2021. “Toward a Principled Bayesian Workflow in Cognitive Science.” *Psychological Methods* 26 (1): 103–26. <https://doi.org/10.1037/met0000275>.

1. Prior predictive checks
2. Fitting the brms model
3. Posterior predictive checks
4. Stability of Bayes factors against MCMC draws

Next, we illustrate these four steps with the model on STAIT-5 as dependent variable, and with *AICT Training Duration* as main variable of interest.

### 1. **Prior Predictive Checks**

As a first step in the Bayesian workflow, we conducted prior predictive checks to assess the plausibility of the prior distributions before fitting the model to the observed data. The outcome variable, *STAIT-5*, was a rescaled version of the original STAI anxiety score, ranging between 0 and 1 (original scale: 5–20).

We specified a weakly informative prior on the intercept, centred on the empirical mean of the STAIT-5 scores (i.e., normal (0.30, 0.05)), and a prior centred at zero for the regression coefficients (normal (0, 0.05)), reflecting no strong assumptions about the direction or magnitude of predictor effects.

Figure 4 shows the results of the prior predictive simulation based on 500 draws. The top panel displays quantile bands of the simulated outcome values across all draws. The middle panel shows the distribution of the mean values of the simulated outcomes, with reference lines indicating the empirical minimum, maximum, and median from the observed data. The bottom panel shows the distribution of simulated standard deviations, overlaid with the empirical standard deviation.

These checks indicate that the prior distributions yield predictions that are consistent with plausible STAIT-5 values. No adjustments to the priors were necessary at this stage.

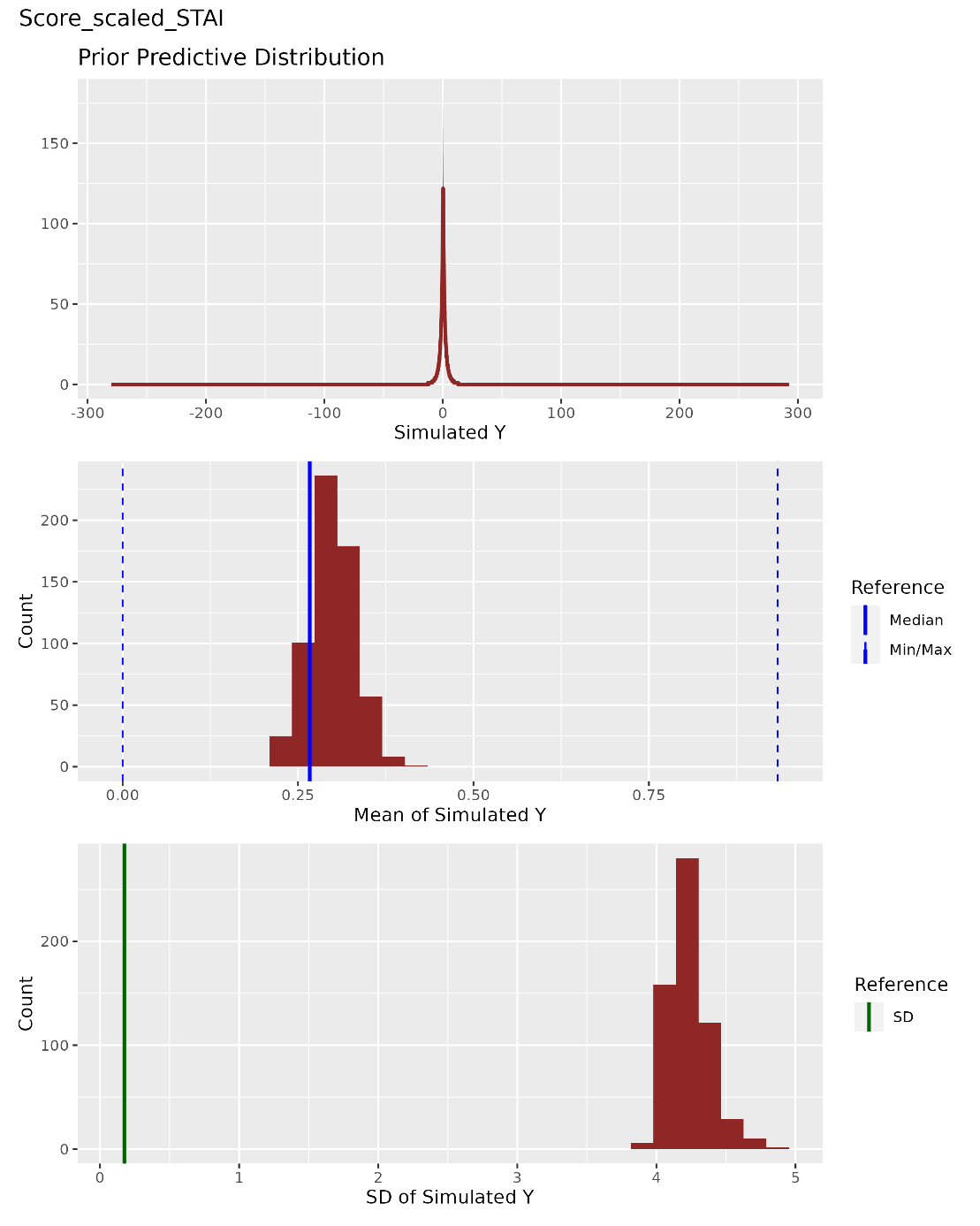

**Supplementary figure 4. Prior predictive checks for the STAIT-5 model.** The top panel displays quantile bands (10–90%, 20–80%, 30–70%, and 40–60%) of simulated outcome values (*STAIT-5*) from the prior predictive distribution. The middle panel shows the distribution of means across simulated datasets, with vertical lines indicating the minimum, maximum, and median of the observed data (blue). The bottom panel presents the distribution of standard deviations in simulated datasets, with a green vertical line marking the observed standard deviation. Overall, the simulations suggest that the specified priors generate plausible values for the STAIT-5 outcome.

### 2. **Model Fitting**

The STAIT-5 model was fitted using the brms package (Bürkner, 2017) with a Gaussian likelihood. The dependent variable was *Score_scaled_STAI*, scaled to fall within [0, 1]. The model included the following predictors: *Age_scaled, Gender, IPSE, Pratique_AutoHypnose, Exp_Trauma, Exp_Transe_Spontanee*, and the interaction *Age_scaled * Anciennete_Formation_AICT_scaled*.

Based on the prior predictive checks (see previous section), weakly informative priors were used: a normal prior centred at 0.30 with standard deviation 0.05 for the intercept, and a normal prior centred at 0 with standard deviation 0.05 for all regression coefficients.

The model was run using 4 chains, with 43,000 iterations each (2,000 warm-up), and sample_prior = "yes" to enable posterior and prior-based draws. Convergence diagnostics (see Table A1) indicated satisfactory model convergence: all Rhat values were below 1.01, effective sample sizes were large, and no divergent transitions were detected. The adapt_delta parameter was set to 0.9, and the maximum tree depth to 13, to ensure stable Hamiltonian Monte Carlo sampling.

|  | **Estimate** | **Est.Error** | **l-95% CI** | **u-95% CI** | **Rhat** | **Bulk_ESS** | **Tail_ESS** |
| --- | --- | --- | --- | --- | --- | --- | --- |
| *Intercept* | 0.13 | 0.08 | -0.03 | 0.29 | 1.00 | 156913 | 133958 |
| *Age_scaled* | -0.05 | 0.01 | -0.07 | -0.02 | 1.00 | 166123 | 121504 |
| *GenreHomme* | -0.03 | 0.02 | -0.06 | 0.00 | 1.00 | 167192 | 122767 |
| *GenreNonbinaire* | 0.01 | 0.04 | -0.07 | 0.09 | 1.00 | 175391 | 121425 |
| *IPSE* | 0.00 | 0.00 | -0.00 | 0.00 | 1.00 | 164389 | 135820 |
| *Anciennete_Formation_AICT_scaled* | -0.02 | 0.01 | -0.03 | -0.00 | 1.00 | 186525 | 130905 |
| *Pratique_AutoHypnose* | 0.04 | 0.02 | 0.00 | 0.09 | 1.00 | 122993 | 120729 |
| *Pratique_AutoHypnoseOui* | -0.01 | 0.02 | -0.06 | 0.04 | 1.00 | 121855 | 120391 |
| *Exp_TraumaJenesaispas* | 0.03 | 0.02 | -0.01 | 0.08 | 1.00 | 160697 | 126921 |
| *Exp_TraumaJepréfèrenepasrépondre* | -0.02 | 0.04 | -0.10 | 0.07 | 1.00 | 172704 | 120162 |
| *Exp_TraumaOui* | 0.04 | 0.01 | 0.01 | 0.07 | 1.00 | 163488 | 124280 |
| *Exp_Transe_SpontaneeJenesaispas* | -0.02 | 0.02 | -0.05 | 0.02 | 1.00 | 147010 | 126189 |
| *Exp_Transe_SpontaneeOui* | -0.03 | 0.02 | -0.06 | 0.00 | 1.00 | 144988 | 123200 |
| *Age_scaled:Anciennete_Formation_AICT_scaled* | 0.01 | 0.01 | -0.00 | 0.03 | 1.00 | 177398 | 125561 |

**Table A1. Summary of the fitted STAIT-5 model (posterior estimates and diagnostics).** For each parameter, Bulk_ESS and Tail_ESS are effective sample size measures, and Rhat is the potential scale reduction factor on split chains. Rhat values near 1 indicate convergence; effective sample sizes (ESS) quantify sampling precision.

### 3. Posterior Predictive Checks

To evaluate the adequacy of the fitted Bayesian models, posterior predictive checks were conducted using 500 draws from the posterior distribution via the posterior_predict() function. Simulated responses were compared to the observed data using a combination of graphical diagnostics. For each model, we computed predictive histograms by binning simulated outcomes and summarizing them across quantiles (10–90%, 20–80%, 30–70%, and 40–60%). These predictive intervals were visualized as overlapping ribbons and compared directly to the histogram of the observed variable. Additionally, the distribution of posterior predictive means and standard deviations was examined across draws, and compared to the empirical mean, standard deviation, minimum, and maximum of the observed data. These checks allowed us to assess how well each model captured the central tendency and dispersion of observed outcomes. In all cases, the posterior predictive distributions aligned well with the observed data, especially in terms of mean and standard deviation, suggesting a satisfactory fit of the models to the data.

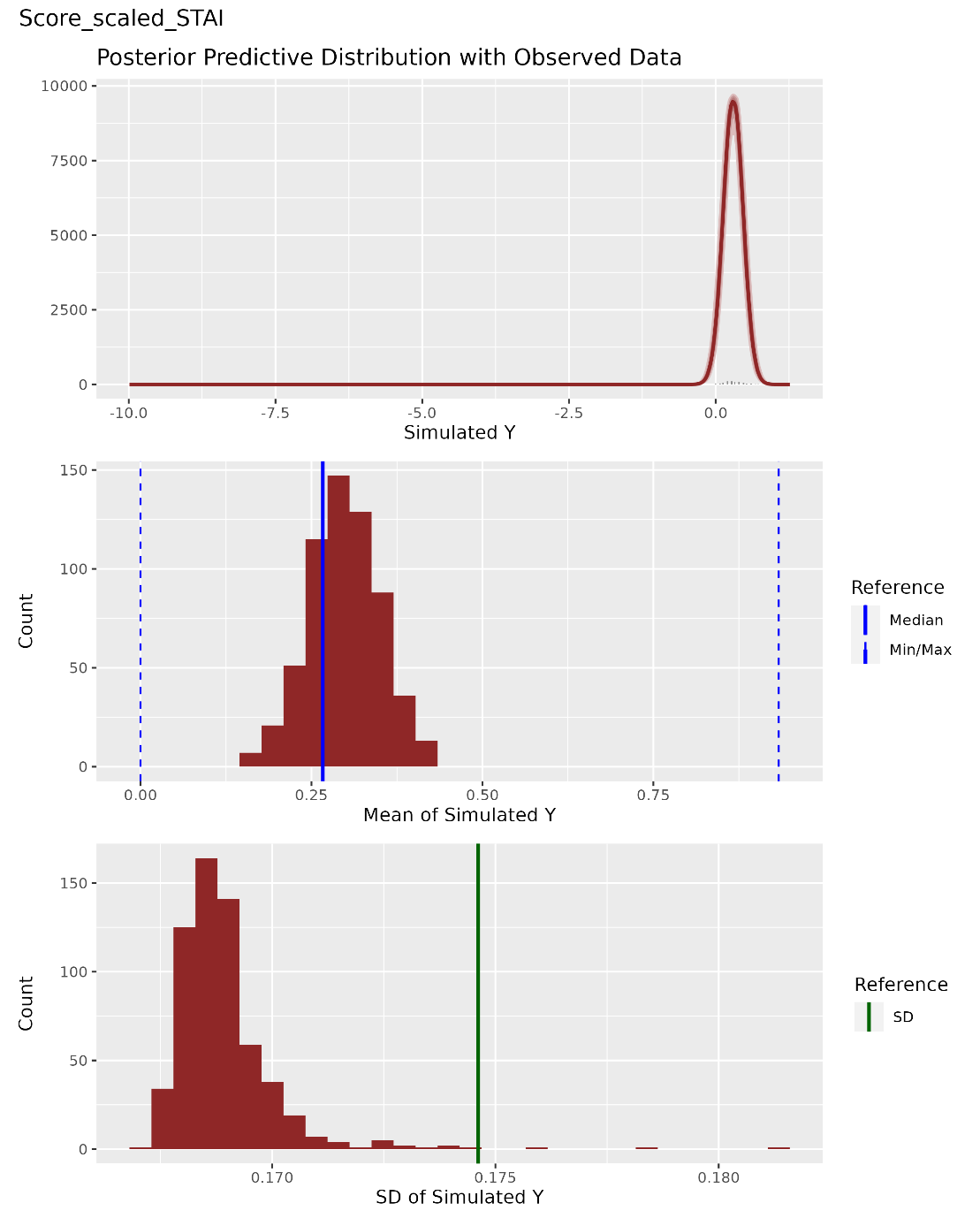

**Supplementary figure 5. Posterior predictive check for the STAIT-5 model.** The top panel shows the observed histogram (in black) overlaid with posterior predictive quantile ribbons. The middle panel displays the distribution of the simulated means, with vertical lines representing the median, min, and max of the observed data. The bottom panel shows the distribution of posterior simulated standard deviations, with a vertical line for the observed SD.

### 4. Stability of Bayes factors against MCMC draws

To assess the robustness of the Bayes Factors supporting the inclusion of our key predictor (*Anciennete_Formation_AICT_scaled*), we conducted stability analyses by refitting each reference model and its corresponding null model (excluding the predictor of interest) across four independent sampling runs. For each model pair, marginal likelihoods were estimated using bridge sampling via the bridge_sampler() function, and Bayes Factors were computed using the bayes_factor() function from the *bridgesampling* package. Previously computed results were loaded when available to avoid redundant computations.

Each sampling run used identical settings: iter = 43000, warmup = 2000, chains = 4, adapt_delta = 0.9, and max_treedepth = 13, with all posterior draws generated without sampling from the prior. No visual figures are presented in this section; instead, the Bayes Factors were stored and compared numerically across runs.

The Bayes Factors proved remarkably stable across repeated estimations, with negligible variability between runs, providing strong support for the reliability of the evidence regarding the inclusion (or exclusion) of the predictor in each model.

### Posterior Inference on Model Parameters: Directional Probability (pd)

To complement the Bayes Factor comparisons conducted on the primary predictors of interest, we computed the posterior *probability of direction* (pd) for all fixed effects in each model. The *pd* represents the proportion of posterior samples that fall on the same side of zero as the median estimate and is considered a Bayesian analogue to frequentist p-values. This metric provides an interpretable measure of effect evidence while remaining compatible with Bayesian principles.

Posterior distributions were extracted using the describe_posterior() function from the *bayestestR* package, with a 95% credible interval and the p_direction statistic enabled. Only fixed effects (parameters prefixed with b_) were retained for analysis. For improved interpretability, parameter estimates were transformed back to their original (unscaled) metrics using the unscale_results() function, based on the original data's means, standard deviations, and observed score ranges.

While Bayes Factors offer strong model-level comparisons, they are computationally intensive and can become impractical when applied exhaustively across all model parameters—especially for control covariates not central to the study's hypotheses. Therefore, we restricted the use of Bayes Factors to focal hypotheses and relied on *pd* values for inference on secondary or control parameters. This hybrid approach offers a robust and efficient framework for Bayesian inference in complex models.
